# Seasonal patterns of dengue incidence in Thailand across the urban-rural gradient

**DOI:** 10.1101/2020.11.25.20186056

**Authors:** Qifang Bi, Derek AT Cummings, Nicholas G. Reich, Lindsay T. Keegan, Joshua Kaminsky, Henrik Salje, Hannah Clapham, Pawinee Doung-ngern, Sopon Iamsirithaworn, Justin Lessler

## Abstract

In Southeast Asia, endemic dengue follows strong spatio-temporal patterns with major epidemics occurring every 2-5 years. However, important spatio-temporal variation in seasonal dengue epidemics remains poorly understood. Using 13 years (2003-2015) of dengue surveillance data from 926 districts in Thailand and wavelet analysis, we show that rural epidemics lead urban epidemics within a dengue season, both nationally and within health regions. However, local dengue fade-outs are more likely in rural areas than in urban areas during the off season, suggesting rural areas are not the source of viral dispersion. Simple dynamic models show that stronger seasonal forcing in rural areas could explain the inconsistency between earlier rural epidemics and dengue “over wintering” in urban areas. These results add important nuance to earlier work showing the importance of urban areas in driving multi-annual patterns of dengue incidence in Thailand. Feedback between geographically linked locations with markedly different ecology is key to explaining full disease dynamics across urban-rural gradient.

## Main Text

Dengue virus (DENV) is an arbovirus estimated to cause 50-100 million symptomatic cases yearly ^1^. Clinical manifestations of DENV infection range from mild dengue fever (DF) to the potentially life-threatening dengue hemorrhagic fever (DHF) and dengue shock syndrome (DSS). DENV has four serotypes, and secondary infection with a different serotype increases the risk of developing severe dengue. DENV was first identified in Thailand in 1949, and all four serotypes have been endemic in the country since at least the 1960s^4^. Within Thailand, there is evidence that, at multi-annual scales, changes in dengue incidence in central Thailand, including Bangkok, precede changes in other parts of the country; possibly driven by population size^3^ or climatic patterns ^2,3^. This pattern of disease emanating from locations with higher mean temperatures and lower average rainfall is also present in multiple countries in southeast Asia^1^. In contrast with these multi-annual dynamics, studies in Cambodia and Vietnam suggest that rural epidemics precede urban epidemics within a dengue season^.5-7^ However, these intra-annual dynamics have not previously been well characterized in Thailand due to limited dengue data at appropriate spatial scales.

The Thai Ministry of Public Health began collecting district-level dengue incidence data across Thailand (77 provinces with 926 districts) in 2003. From 2003 to 2015 there were 1,164,278 reported dengue cases, including 637,807 cases of dengue hemorrhagic fever (DHF) and dengue shock syndrome (DSS) (Fig. 1A). We analyzed these 13 years of combined DHF/DSS incidence data to better understand DENV at multiple spatial and temporal scales within the country. We focus on DHF/DSS because DHF/DSS surveillance is believed to be less sensitive to secular trends in reporting, due to disease severity and clear clinical presentation ^8,9^.

**Figure 1.**
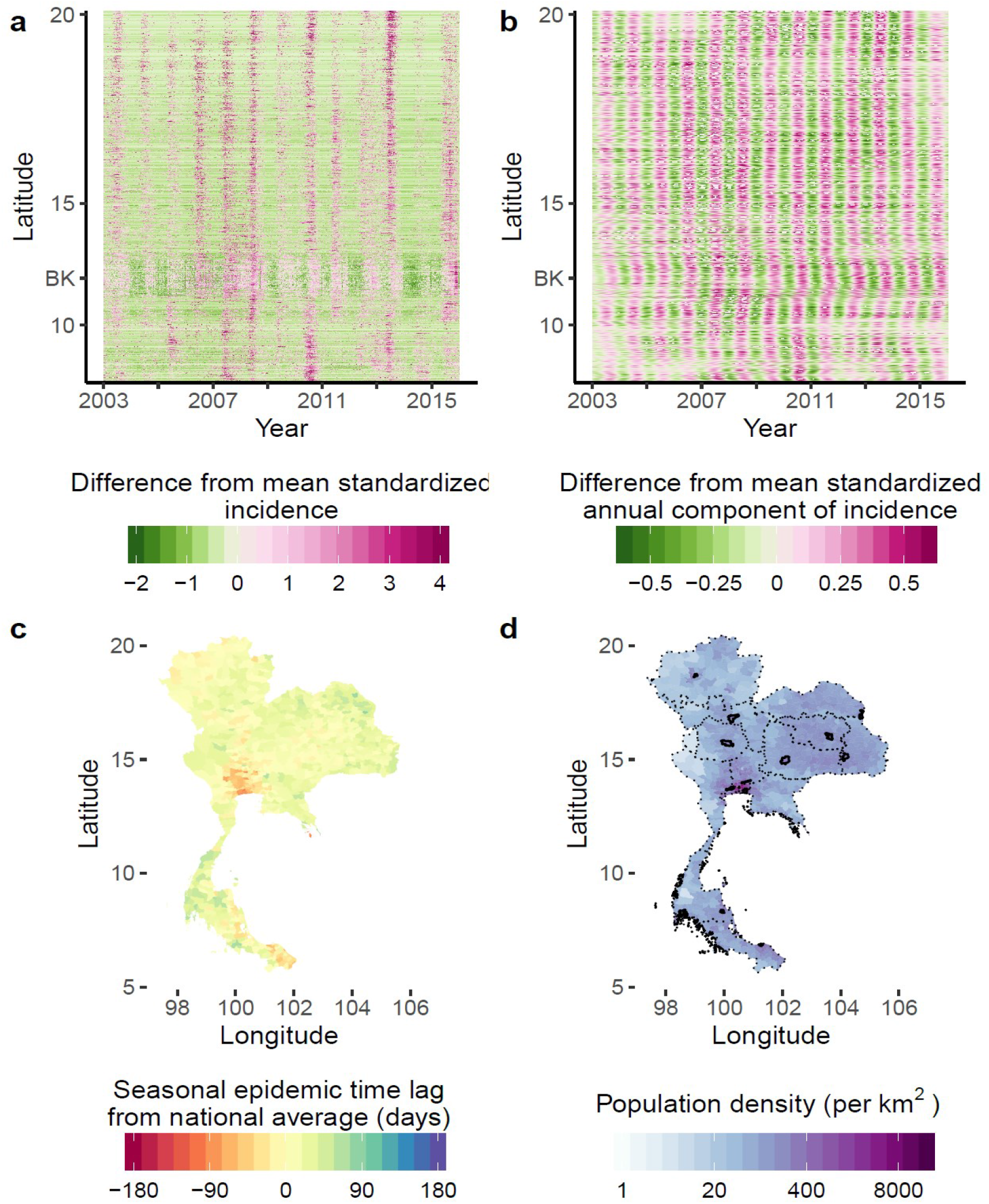
Timing of dengue incidence by geographic location. **a**, Normalized DHF/DSS incidence from 2003 through 2015 for each Thai district ordered by latitude (0.63% of normalized incidence was above 4 and was given the color representing normalized incidence of 4). **b**, Annual component of standardized DHF/DSS incidence for each Thai district ordered by latitude. Each horizontal line in b and c represents a time series of one district. BK represents Bangkok. **c**, Seasonal epidemic time lag of intra-annual DHF/DSS waves of each district relative to the national average. Positive numbers indicate the number of days the intra-annual DHF/DSS waves in a district preceded the aggregated country-level waves, whereas negative numbers indicate a lag. **d**, District level population density. Dotted lines outline borders of each health region. Solid lines outline the urban center of each health region.

Using wavelet analysis, we isolated the annual component (9-14 months) of the normalized, square-root-transformed, time series of DHF/DSS incidence per 100,000 people for each Thai district ^10,11^. To characterize the time lag of seasonal epidemics, we then calculated the phase difference for each district relative to the aggregate country-level time series. We found that within a year, dengue epidemics in more densely populated urban districts such as Bangkok lagged behind the rest of the country (Fig. 1B and 1C). Specifically, seasonal epidemics lagged by 8.9 days (95% CI: 8.0-9.8 days) for each two-fold increase in population density (Fig. 2A). This phenomenon exists locally across 11 of the 13 health regions in Thailand (IQR 1.3-8.5 days) and all 6 meteorological regions (IQR 2.9-8.9 days) (Extended Data Fig. 1, Extended Data Table 1).

**Figure 2.**
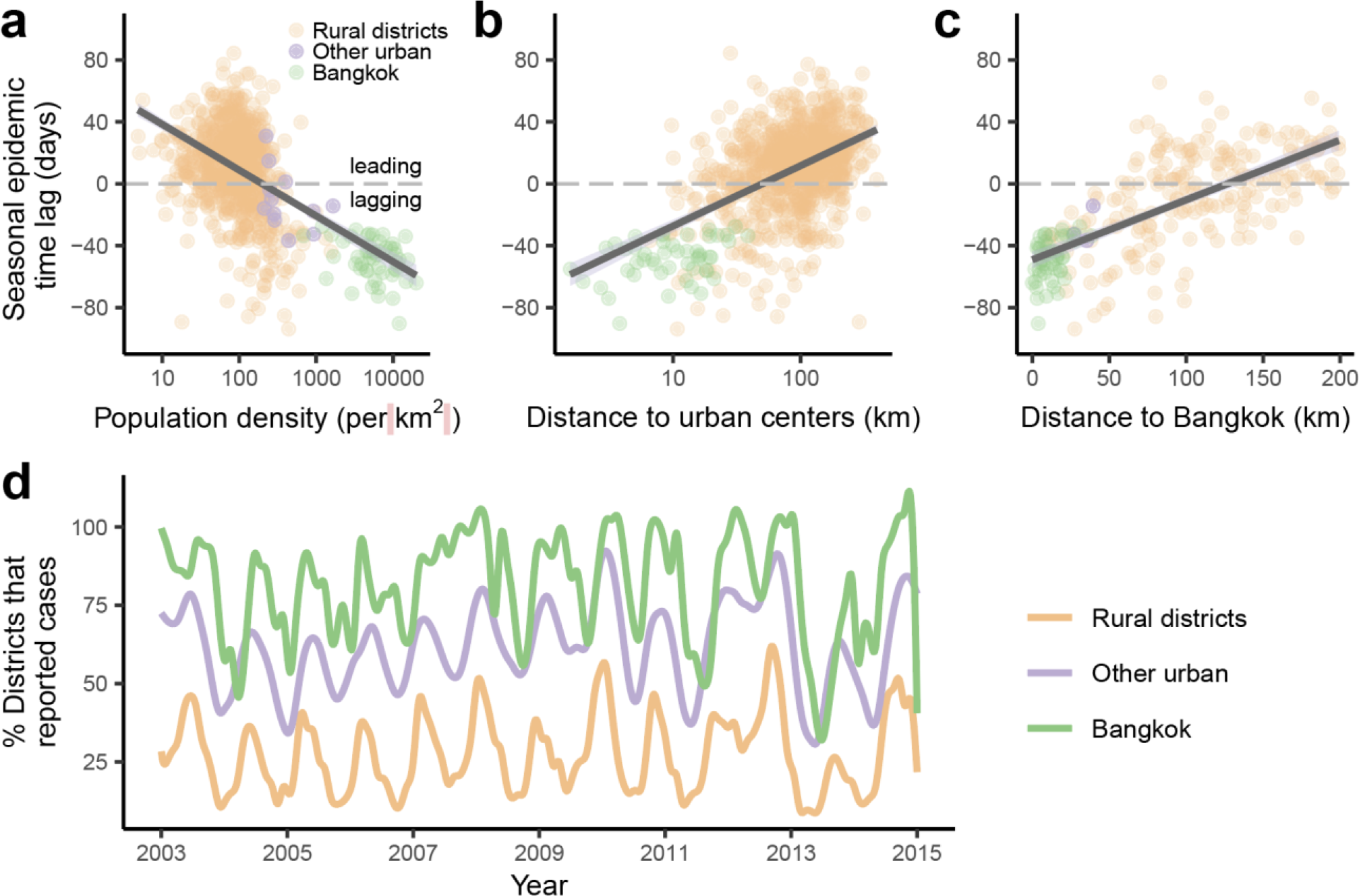
Characteristics associated with timing of dengue seasonal epidemic in Thailand. Association of seasonal epidemic time lag with (**a**) the population density of a district, (**b**) the distance between a district and its regional urban center, defined as the district with the highest population density in a health region, and (**c**) the distance between central Bangkok and a district within a 200km radius from central Bangkok. The seasonal epidemic time lag is defined as the number of days the intra-annual DHF/DSS waves in a district preceded (positive numbers) or lagged (negative numbers) the aggregated country-level DHF/DSS waves. Each point represents a district, and colors represent 3 types of districts, districts in Bangkok (green), other urban districts (defined as the districts with the highest population density in each of the 11 health regions, not including Bangkok; purple), and all other districts (orange). **d**. Proportion of districts that reported DHF/DSS cases over the study period, by the 3 types of districts described above.

The timing of dengue epidemics was also highly associated with distance from major urban centers. Seasonal epidemics shifted ahead by 16.7 days (95%CI: 14.3 - 19.1 days) for each 100km increase in distance from a regional urban center, defined as the district with the highest population density in a health region (Fig. 2B). In 12 of the 13 health regions, this trend was present relative to the regional urban center (Extended Data Fig. 2, Extended Data Table. 1). The trend was most pronounced within a 200km radius from central Bangkok, the capital of Thailand and the region with the highest population density, where epidemics shifted early by 3.9 days (95%CI: 3.3-4.4 days) for each additional 10km of distance from central Bangkok (Fig. 2C). We found that distance to Bangkok appears to be the main driver of the timing of seasonal epidemics in the health regions adjacent to Bangkok (health regions 4,5,6), but in the more remote regions of Thailand distance to regional urban centers appears to be the main influence (Extended Data Table 2).

Seasonal epidemics in districts with sporadic case counts may appear to precede urban areas based on wavelet analysis even though epidemics start later compared to the urban areas. We excluded the districts where we see this type of discrepancy between incidence patterns and results from wavelet analysis, and we found that the association between phase difference and ruralness of a district still holds around Bangkok and Southern region of Thailand where sample size remained adequate (Extended Data Fig. 3). We also suspected that urban areas may disproportionately influence the effect size for the regressions since districts tend to be much smaller in urban areas. To address this, we aggregated districts to a standardized size and found that the associations were maintained (Extended Data Fig. 4). The observed association is also robust to analysis based on dengue fever incidence (rather than only DHF/DSS incidence) (Extended Data Fig. 5).

**Figure 3.**
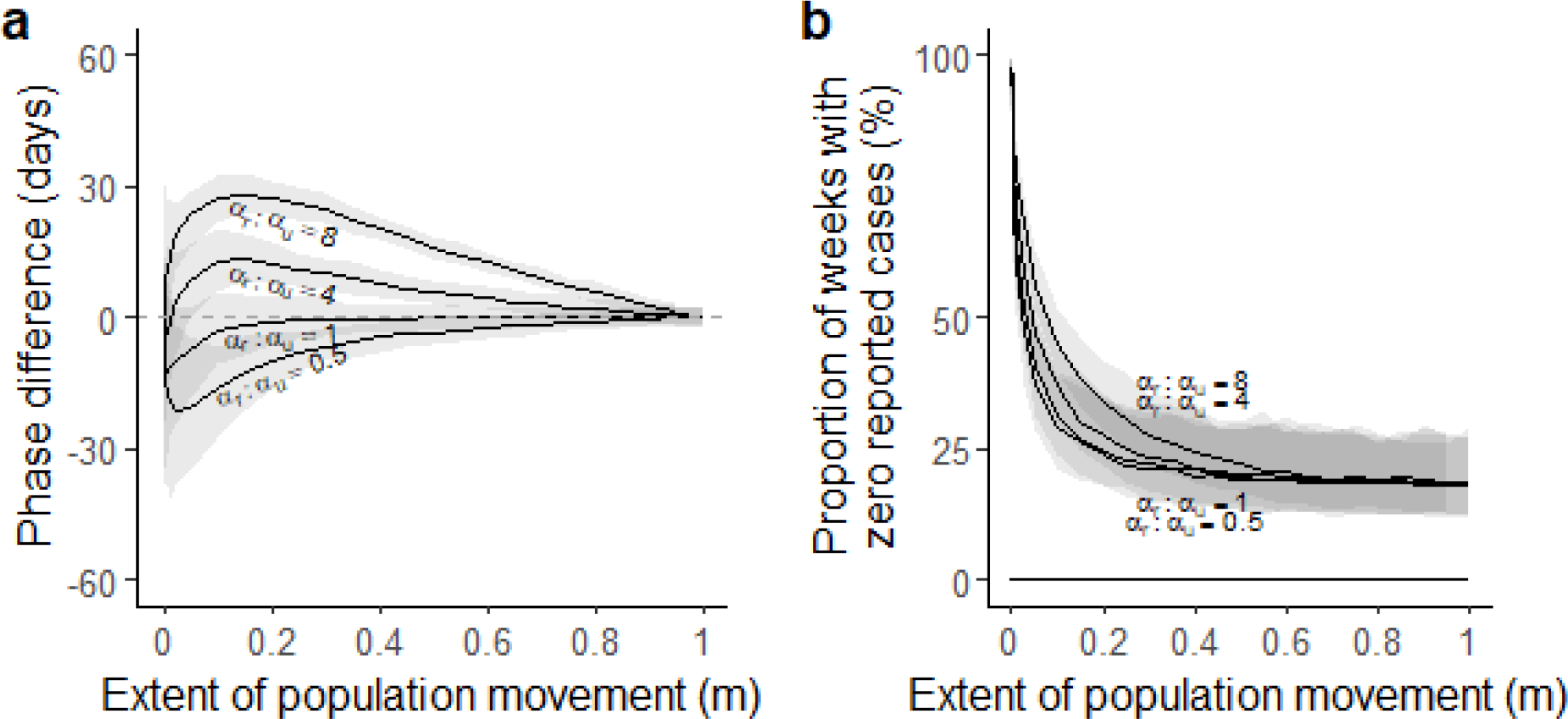
Relationship between timing of dengue seasonal epidemics, fadeout, and seasonal forcing in a simulated two-patch model. **a**, Stochastic phase differences of seasonal epidemics on annual scale between the urban and rural patch for various parameter combinations. *α*_*u*_= 0.2, α_*r*_= 0.1, 0.2, 0.4, 0.8, and 0<m<1; Positive numbers indicate the number of days the intra-annual waves in the rural district preceded that in the urban district, whereas negative numbers indicate a lag. We ran 50 simulations for each combination of parameters for up to 60 years and calculated average phase difference between the simulated seasonal epidemics (0.8 - 1.2 years) in the urban and rural patch. Black curves represent the smoothed median value of the average phase differences. Shades represent the 95% confidence interval. **b**, the proportion of weeks without cases in simulated urban (all close to zero) and rural (solid curves) epidemics for the same set of parameters.

Since population density tends to decrease with distance from major urban centers, we questioned whether the two measures of ruralness had independent effects on the timing of seasonal epidemics. Bivariate models showed a substantial improvement in fit over both univariate models (AIC improvement of 176 vs. distance only model, and 149 vs. density only), and the bivariate models showed a significant effect of both distance and density, though the magnitude of both was decreased (8.1 days [95% CI: 5.6-10.6] per 100km distance, 7.3 days [95% CI: 5.8-8.8] per two-fold increase in density). Results were similar within health regions (Extended Data Table 1).

Though rural dengue epidemics precede urban epidemics within a season, it does not appear that dengue virus circulation is maintained in rural areas during the off-season, acting as a seasonal reservoir for urban areas. Rather, we found that local dengue fade-outs were less likely in Bangkok and other urban centers than in the rest of Thailand (Bangkok is significantly more likely (p<0.05) to report a case than rural district over 98% of the study period; over 60% when comparing other urban district with rural districts, Fig. 2D). The frequency of fade-outs, defined as weeks with no reported cases, was associated with both population density and distance from urban centers (Extended Data Fig. 6B, 6C); and these differences were greater than would be expected based on sampling effects alone (i.e., they exceed the difference in the frequency of observed zeros expected due to sampling from proportionally fewer cases in rural areas, Extended Data Fig. 6A). We think these differences are unlikely the result of differences in reporting rates between districts, because Thailand has universal healthcare and DHF and DSS are both clinically severe manifestations of DENV.

To explore the mechanisms that could drive rural epidemics to precede urban epidemics, and understand whether local fade-outs were theoretically inconsistent with this phenomenon, we created a simple two-patch metapopulation model for a single strain pathogen (See Methods for a detailed description of the model). We hypothesized that a stronger amplitude of seasonal forcing in the rural patch would lead to earlier seasonal epidemics in that patch, even if the frequency and phase of seasonal drivers of transmission were otherwise identical. We further hypothesized that, in a discrete, stochastic version of this model, fade-outs would be frequent in the rural patch but seasonal epidemics in the rural patch would continue to precede the urban epidemic.

Our simulations bore out these hypotheses. In both weakly and strongly linked communities, yearly epidemics in the rural patch precede epidemics in the urban patch when the amplitude of seasonal forcing was larger in the rural patch, with equal baseline transmission rates (Fig. 3A, Extended Data Fig. 7). This time-lag increases as the difference in the amplitude of seasonal forcing of the two patches increases. However, in unlinked or fully linked communities, this time-lag does not exist regardless of the difference in amplitude of seasonal forcing. For all combinations of seasonal forcing and mixing parameters, local disease fade-outs (i.e. proportion of weeks without cases) are more frequent in the rural patch compared to the urban patch (Fig. 3B, Extended Data Fig. 7). We further showed that it was possible to have the rural patch lag the urban patch at multi-annual scales, while still having rural epidemics precede urban epidemics within dengue seasons (Extended Data Fig. 8), thus showing that our findings are theoretically consistent with previous work on multi-annual cycles.^3^

Plausible drivers of higher seasonal forcing in rural Thai districts exist. For instance, seasonal variance in the Normalized Difference Vegetation Index (NDVI) and average daytime land surface temperature is highly correlated with population density in Thailand. We found that the variance of NDVI decreased by 0.27 standard deviation from the mean variance (95%CI: 0.22-0.32; z-score of the variance of NDVI) for each two-fold increase in population density in areas outside of tropical or temperate rainforests (Extended Data Fig. 9 and 10). Variance of land surface temperatures decreased by 0.33 (95%CI: 0.29-0.38) standard deviation from the mean for each two-fold increase in population density, after adjusting for the mean (Extended Data Fig. 11 and 12). Although not completely explaining the impact of ruralness on timing of seasonal epidemics, both NDVI and land surface temperature have strong associations with timing of dengue seasonal epidemics even after adjusting for population density (Extended Data Table 5). Even though total rainfall variance is not associated with local population density (Extended Data Fig. 13), seasonal variation in surface water appears to be smaller in urban environments compared with rural areas, because the larger impervious surface and more water-filled artificial containers in urban environment retains water longer. These seasonal difference in environmental drivers was thought to be reflected in seasonality of *Aedes* population ^12–15^. Likewise, rural areas of Thailand are largely agricultural, with greater seasonal differences in human behaviour compared to urban areas, a phenomena linked to measles seasonality in countries like India and Niger ^16-19^. Variance of environmental factors is an oversimplified representation of seasonal forcing, and more work is needed to tease apart the effect of seasonal human behavior, while accounting for the difference in immunity profile, in order to identify drivers of seasonal forcing.

In this study, we showed that epidemics in rural areas precede urban ones within a year in Thailand, a phenomena previously masked due to a lack of surveillance data at fine spatio-temporal resolution. This same data suggest that dengue fadeouts (i.e., local extinctions) are common in rural areas during the off season, consistent with previous studies showing that dengue ‘overwinters’ in larger urban centers such as Bangkok ^4^. These two phenomena on the surface may seem contradictory, and likewise, earlier seasonal epidemics in rural areas may seem at odds with earlier work showing ‘travelling waves’ emanating from Bangkok ^3^. However, simple computational models show that these results are in fact consistent with theory and would largely be explained if seasonal forcing in rural areas was greater than that seen in urban locations. Several possible drivers of stronger seasonal forcing in rural areas exist, but more work is needed to show which of these is in fact driving stronger seasonality. Nevertheless, our findings have important practical and scientific implications. The knowledge that dengue epidemics in rural areas are starting several weeks, or more, before those in populous urban areas suggests that improved, timely, dengue surveillance in those areas may provide important early information about the nature of each year’s epidemics possibly improving early warning systems. From a scientific standpoint, this work shows that we must be careful about generalizing timings of epidemics to the actual movement of viral lineages and vice versa. Both empirical evidence and theory show that, at the start of a dengue season, viral lineages may move from urban to rural areas, where stronger seasonal forcing may accelerate the epidemic. This leads to an earlier explosion of cases in rural areas and in doing so provide an additional seasonal driver for the urban epidemic. Hence, to understand the full dynamics of diseases in geographically linked systems, we must not think simply in terms of sources and sinks. Instead we should attempt to understand how feedback loops between areas with markedly different disease ecology can lead to the phenomena we observe.

## Data Availability

Data can be made available upon request.

## Materials and Methods

### Dengue data and study settings

We obtained daily district level dengue case counts (including DF, DHF, and DSS cases) collected by the Thai Ministry of Public Health (MOPH) from January 1^st^ 2003 to December 31^st^ 2015. We aggregated daily time series data into weekly incidence data at the district level. We then square-root-transformed and normalized the weekly incidence to have a mean of zero. We obtained 2013 district-level population data internally from the Thai Ministry of Interior. We assumed that the population remained constant over the course of our study period, since changes in population size and, as a result, dengue incidence do not change the estimates of phase angles.

We obtained and adapted the district level administrative boundaries of Thailand from the GADM database (http://www.gadm.org/download). District designation/boundaries were kept consistent over the study period. Our data included 77 Thai provinces and 926 Thai districts. The area of each district was determined based on the modified GADM shapefile. We obtained the division of 13 health regions from the Thai MOPH. Division of the 6 geographic regions and 6 meteorological regions were released by the Thai National Research Council.

### Wavelet transformation

We used wavelet analysis to isolate the intra-annual component of the normalized time series of dengue incidence, for periods between 0.8 to 1.2 years. Detailed methods for wavelet analysis have been previously described ^10,11^. The Morlet wavelet *ψ*is a localized periodic function, ideal for analyzing periodic behavior such as seasonal dengue incidence patterns. We computed Morlet wave transforms for each district using a nondimensional frequency *ω*_0_=6and a periodicity step size δj of 0.25 ^10,11^, similar to previous analyses.

The continuous wavelet transform is the convolution of time series *x*_*k*_, whose observations are equally spaced with time interval *δt*, and the wavelet *ψ*^∗^at time *t* and scale (or periodicity) *s*. For each scale *s* and time interval *δt*, the continuous wavelet transform of a time series is defined as:

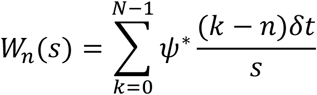

where ψ represents the Morlet wavelet function and * the complex conjugate. n represents the time index, ranging from zero to the total number of time points N. We computed wavelet transforms for scales ranging from 0.8 to 1.2 years using the dplR package in R.

For each district, we then computed the average of wavelet transforms across periodicities in the annual (0.8-1.2 years) band using the scale-averaged wavelet power:

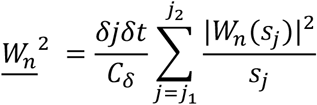

We reconstructed the annual component of the epidemic cycles using:

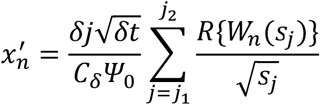

where *δj* was 0.125, *Cδ* was 0.776, and values for *j*1 and *j*2 were 0.8 and 1.2 years.

### Phase difference between time series of dengue incidence

The wavelet transformation encodes the time- and frequency-specific phase. We computed phase angles for annual cycles as indicators of timing of seasonal epidemics, as phase indicates the angular position of each point in its cyclical trajectory. For each district, we computed the average phase angle *ϕ*_*n*_for annual cycles as the circular average of periodicity-specific phase angle *ϕ*_*n*_(*s*)across the annual periodicity bandwidth, 0.8 - 1.2 years.

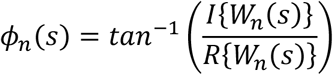

We used these phase angles to calculate a time series of phase difference for each district, with respect to the aggregated national level data ^10,11^. The phase angle difference *θ* was constrained between −*π* and *π*.

To understand the timing of seasonal patterns of dengue incidence and the impact of urban-rural gradient across Thailand, we then averaged the time series of phase difference of each district, and assessed its association with 1) district-level population density, and 2) distance from regional urban centers using univariate linear regression. We used bivariate analyses including both population density and distance to regional urban centers to understand whether the two measures of ruralness had independent effects on the timing of seasonal epidemics. We repeated the same analyses by health region (geographic region, or meteorological region). We defined regional urban centers as districts with the highest population density in each health region (geographic region, or meteorological region). We performed sensitivity analyses to understand the relative impact of distance to Bangkok and distance to regional urban centers on timing of seasonal pattern of dengue incidence.

### Characterize local dengue fade-outs

We explored the association of the frequency of fade-outs with population density and distance from urban centers. In addition, we assessed whether the differences in frequency of fade-outs between districts exceeded what would be expected based on sampling effects alone. We defined frequency of fade-outs as the number of weeks where no cases were reported in a district. We assume the expected number of cases follows a binomial distribution, and thus the expected frequency of fade-outs is defined as *Pr(Binomial(p, AR* × *N*_*i*_)= 0), where *p* represents dengue detection rate, *AR* represents dengue weekly attack rate, and *N*_*i*_represents the population size of a district *i*. We estimated the proportion of district pairs (comparing each district to one with higher population size) whose relative frequency of fade-outs exceeded the expected relative frequency of fade-outs, and assessed its association with the product of detection rate and attack rate, *p* × *AR* (see Extended Data Fig 4). We also assessed how this association varies by difference in population size between district pairs.

### Climate Indices

We explored plausible drivers of higher seasonal forcing in rural Thai districts compared to urban districts. We obtained 1) gridded, 16-day, Normalized Difference Vegetation Index (NDVI) with 0.1*0.1 degree resolution for 2009, 2) gridded, 8-day, average daytime land surface temperature for 2001-2010 with 0.1*0.1 resolution, 3) gridded, daily rainfall with 0.25*0.25 resolution from 2009 (http://neo.sci.gsfc.nasa.gov). We used univariate linear regressions to assess the associations of 1) variance of NDVI and 2) variance of rainfall with population density of the grid cells outside of tropical or temperate rainforests (defined as the grids with mean NDVI over 0.6). We also used bivariate linear regression to assess the association of the variance of average daytime land surface temperature with population density after accounting for the mean of average daytime land surface temperature that is positively associated with its variance.

### Coupled Two-Patch SIR Models with Seasonal Forcing

We created a two-patch metapopulation model for a single strain pathogen to test the hypothesis that stronger seasonal forcing in the rural patch alone would lead to an earlier seasonal epidemic in that patch. We calculated phase difference *θ* of the reconstructed seasonal and multiannual epidemics between the two patches.

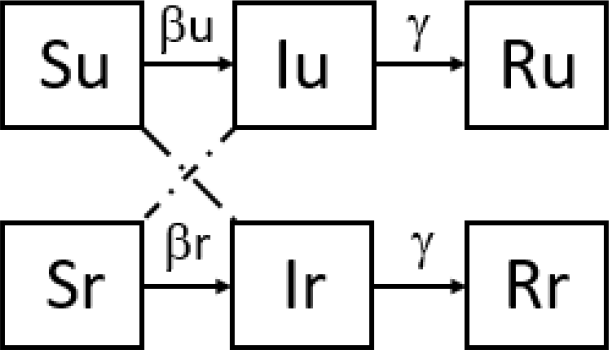

Here, *S*_*u*_, *I*_*u*_, *R*_*u*_, *S*_*r*_, *I*_*r*_,and *R*_*r*_ are the number of susceptibles, infectious, and recovered individuals in the urban and rural patch. To mimic the dynamics of coupled urban and rural districts in Thailand, we set the total population size of the rural district *N*_*r*_ to be 40,000, which approximates the median population size for rural districts in Thailand. We set the population size of the urban district *N*_*u*_ to be 4,000,000, consistent with the critical community size for dengue ^20,21^. In both models, individuals are born susceptible to dengue infection at a birth rate of *μ*=11 per 1,000 people per year ^22^. After infection, individuals recover at a fixed rate *γ* where 1/*γ* represents the infectiousness period, the same for urban and rural patches. We set the death rate equal to the birth rate, for both patches. Seasonality in the rate of transmission is modeled using a simple sinusoidal function, such that *β(t)*= *β*_0_(1 *+* α *⋅ cos(t* − *φ*)), where *β*_0_is the baseline transmission rate, α is the amplitude of seasonality, and ϕ is the seasonal offset parameter. We assume that the baseline transmission rate *β*_0_is 0.25/day. Seasonal forcing terms independently influence the dynamics in each patch. Amplitude of the seasonal forcing terms α could vary by patch though the baseline transmission rates *β*_0_are always the same. 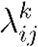 represents the FOI of individuals in the patch i meeting individuals in the patch j in patch k.

The model assumes that contacts between the urban and rural population can occur in either patch and are inversely proportional to the population size of the two patches. *m* indicates the extent of population mixing between the two patches such that *m* = 0indicates no mixing, while *m* = 1 indicates the full mixing scenario under which proportion of population that are in contact with the other patch is perfectly inversely proportional to the population size of the population. That is, when *m* = 1, the proportion of rural population that are in contact with the urban patch is 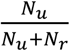, and the proportion of urban population that are in contact with the rural patch is 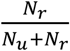, Probabilistic transition between compartments at each time step follows a binomial distribution. Stochastic simulation of each combination of parameters initialized at their deterministic equilibrium.

To show that rural areas can precede urban in intra-annual patterns while lagging on the multi-annual scale, we then updated rate of infection to the sum of two sinusoidal waves, one with 1-year periodicity as described above and the other with 3-year periodicity, such that *W*_*n*_(*t*) = *β*_1_(1 *+* α *⋅ cos(t* − *φ*)*)+ β*_2_(1 *+* α *⋅ cos(t*/3 − *φ*)). The rural sinusoidal waves with 3 year periodicity lags the urban one by 2 years. The parameters used here are *β*_1_ = 0.15, *β*_2_ = 0.05, *φ* = 180, and *m* = 0.1. Amplitude of seasonal forcing of the two patches are α_*u*_ = 0.2 and α_*r*_ = 0.8 respectively.

| Parameter | Biological meaning | Default value or range of values |
| --- | --- | --- |
| $\beta_0$ | Baseline transmission rate | 0.25/day, same for urban and rural districts |
| $\alpha_u, \alpha_r$ | Amplitude of seasonal forcing | 0 to 1 |
| $\phi$ | Seasonal offset parameter | 180 days, same for urban and rural districts |
| $1/\gamma$ | Infectiousness period | 10 days |
| $m$ | Extent of population movement | 0 to 1 |
| $\mu$ | Birth rate and death rate | 11 per 1,000 people per year |
| $N_u$ | Urban population size | 4 million |
| $N_r$ | Rural population size | 40,000 |

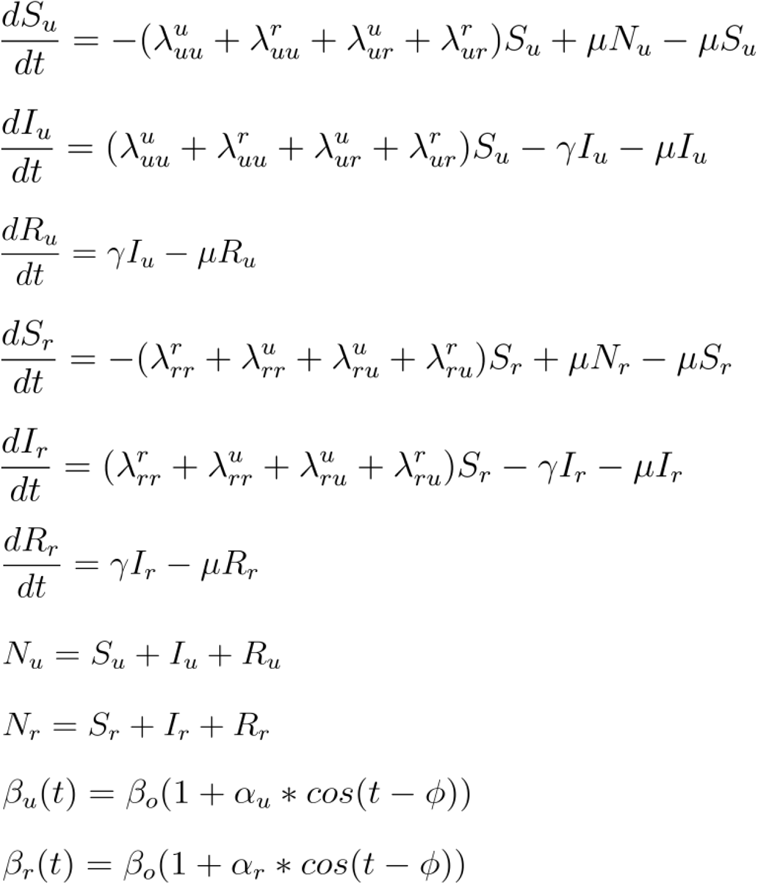

FOI of *S*_*u*_ meeting *I*_*u*_ in the urban patch:

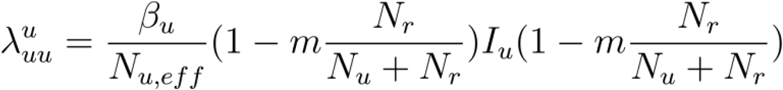

FOI of *S*_*u*_ meeting *I*_*u*_ in the rural patch:

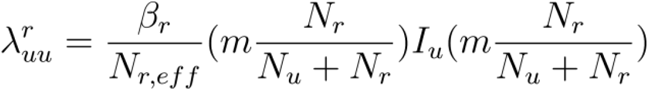

FOI of *S*_*u*_ meeting *I*_*r*_ in the urban patch:

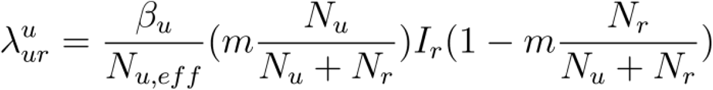

FOI of *S*_*u*_ meeting *I*_*r*_ in the rural patch:

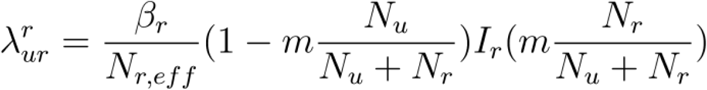

FOI of *S*_*r*_ meeting *I*_*r*_ in the rural patch:

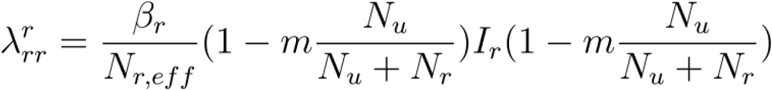

FOI of *S*_*r*_ meeting *I*_*r*_ in the urban patch:

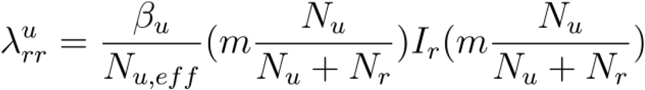

FOI of *S*_*r*_ meeting *I*_*u*_ in the urban patch:

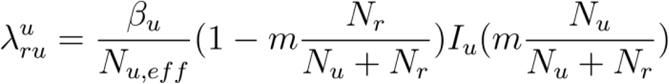

FOI of *S*_*r*_ meeting *I*_*u*_ in the rural patch:

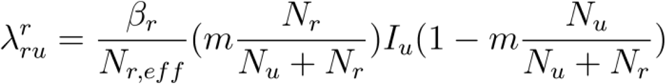

Where 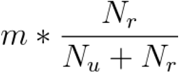 is the probability of urban people traveling to the rural patch,

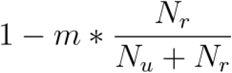 is the probability of urban people staying in the urban patch,

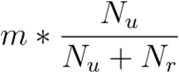 is the probability of rural people traveling to the urban patch, and

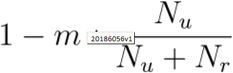 is the probability of rural people staying in the rural patch.

We show that the effective population size in the urban patch at each time point equates to the initial population size in the urban patch. Same for the rural patch.

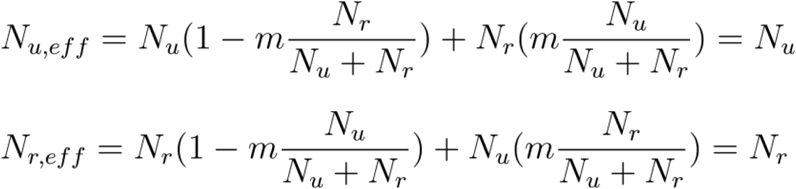

