## Supplemental Material for "Seasonal patterns of dengue incidence in Thailand across the urban-rural gradient"

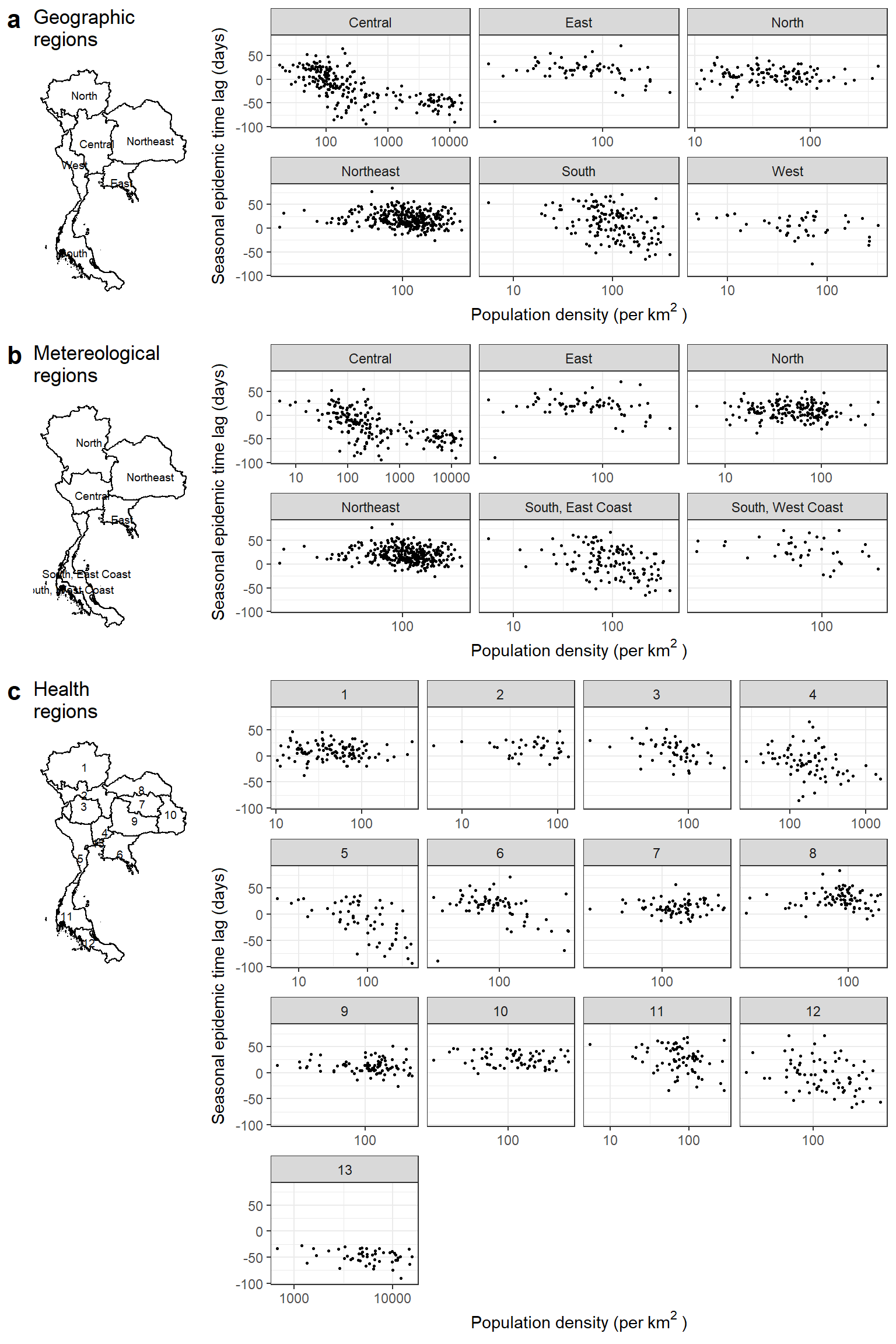

Figure S1: Association between seasonal epidemic time lag and population density by region, a) 6 meteorological regions, b) 6 geographic regions, and c) Bangkok and 12 health regions defined by the Regional Office of Disease, Prevention and Control (ODPC). The seasonal epidemic time lag is defined as the number of days the intra-annual DHF/DSS waves in a district preceded (positive numbers) or lagged (negative numbers) the aggregated country-level DHF/DSS waves.

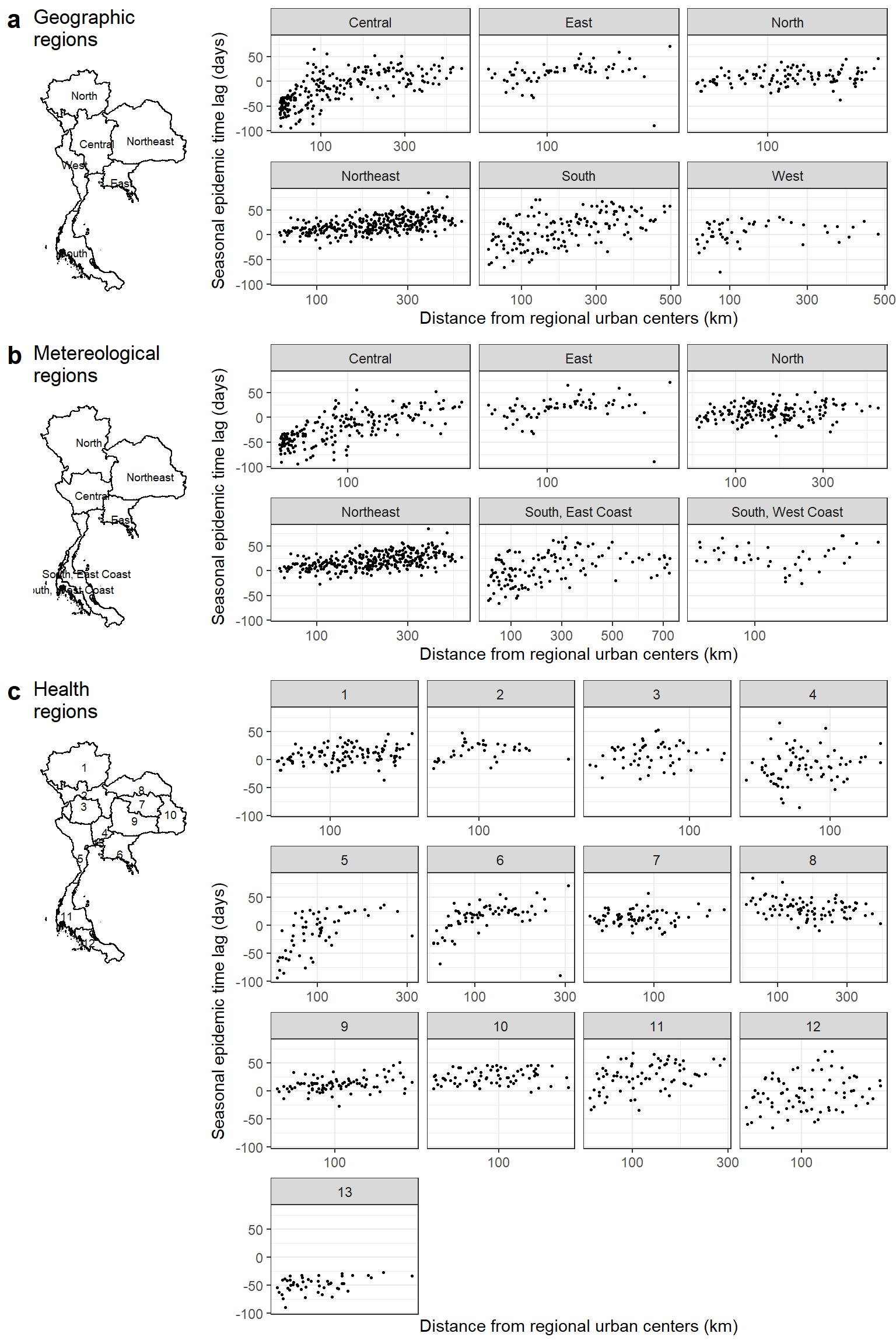

Figure S2: Association between seasonal epidemic time lag and distance from regional urban centers by region, a) 6 meteorological regions, b) 6 geographic regions, and c) Bangkok and 12 health regions defined by the Regional Office of Disease, Prevention and Control (ODPC). The regional urban center in each region is defined as the non-island district with the highest population density. The seasonal epidemic time lag is defined as the number of days the intra-annual DHF/DSS waves in a district preceded (positive numbers) or lagged (negative numbers) the aggregated country-level DHF waves.

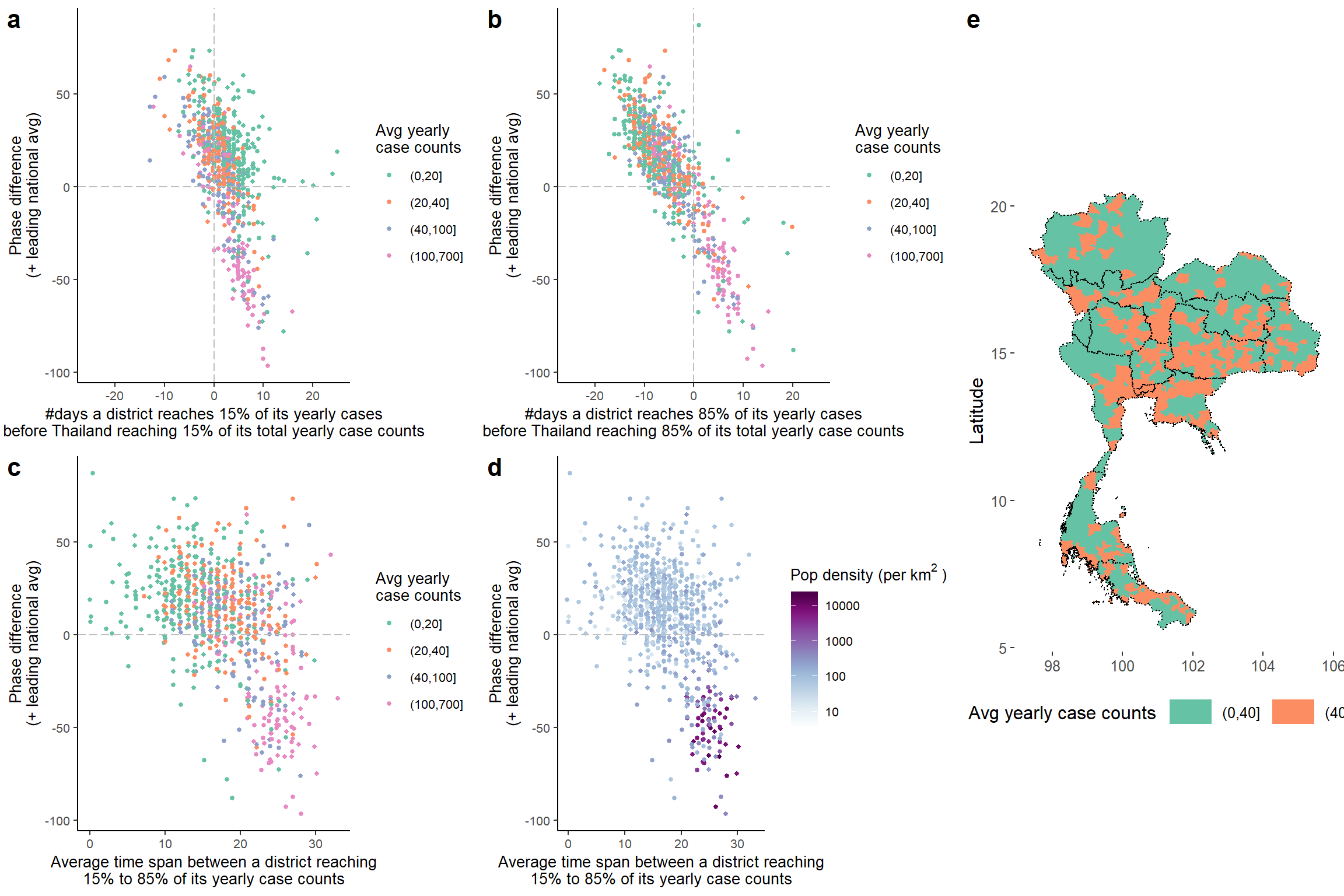

Figure S3: The ability of wavelet analysis to differentiate the true preceding pattern from noise. a,b) Comparison between the start (a) or end (b) of an epidemic and phase difference of a district relative to the national average. Start (end) of an outbreak is represented by the number of days a district reaches 15% (85%) of its yearly case counts before Thailand reaching 15% (85%) of its total yearly case counts. Phase difference shows the number of days the intra-annual DHF/DSS waves in a district preceded (positive numbers) or lagged (negative numbers) the aggregated country-level DHF/DSS waves. Color represents the average yearly case counts of a district. Seasonal pattern of incidence in districts that fall under the second and fourth quadrants truly precedes (or lags) that of the national average, whereas seasonal pattern of incidence in districts that fall under the first and third quadrants represent noise (i.e., sporadic cases). We subsetted districts to those that truly preceded or lagged the national average (i.e., those that fall in the second and fourth quadrant) for sensitivity analyses (regression results shown in Table II). c) Association between phase difference and time span of seasonal epidemics, defined as time from a district reaching 15% to reaching 85% of its yearly case counts. Color represents average yearly case counts of a district. d) is similar to c), but color represents population density of a district. e) The spatial distribution of districts with average case counts above and below 40 cases a year.

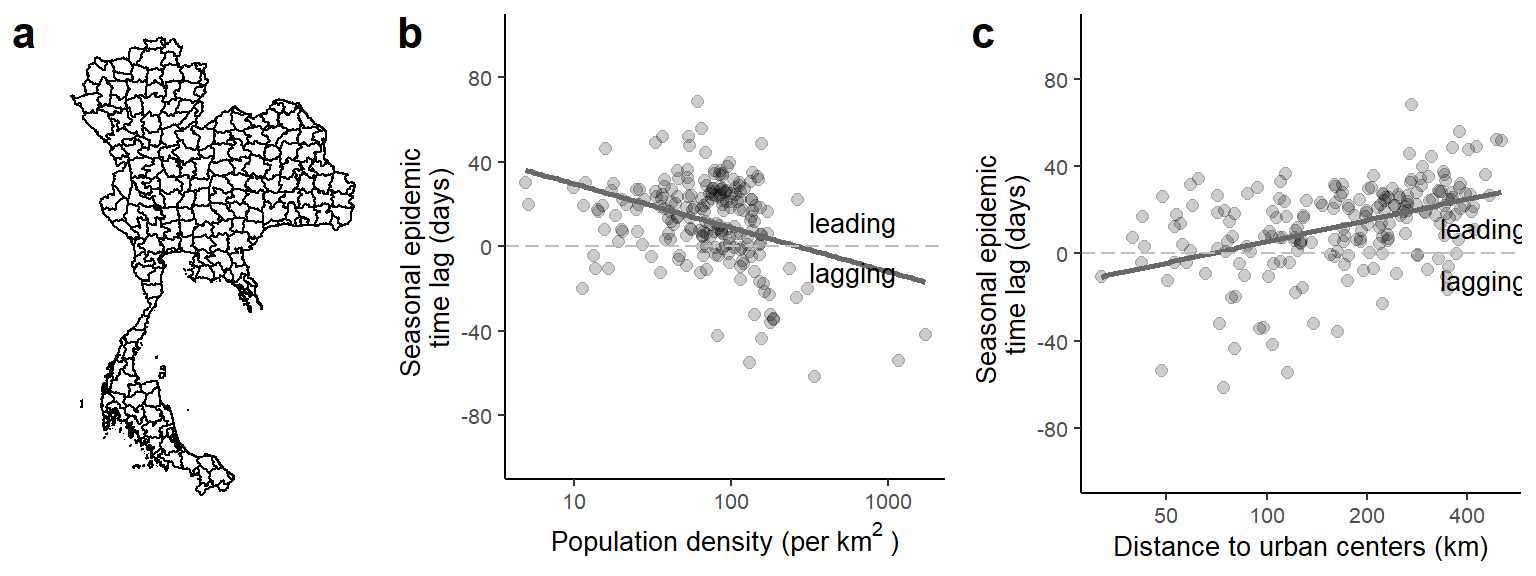

Figure S4: Factors associated with seasonal epidemic time lag after regrouping districts. a) Geographic division of the 209 regrouped districts. The adjacent districts whose centroids fall within the same grid (0.5 degree latitude by 0.5 degree longitude) were combined into one regrouped ‘district’. b) The association between timing of seasonal epidemic and population density of a regrouped district. The seasonal epidemic time lag is defined as the number of days the intra-annual DHF/DSS waves in a regrouped district preceded (positive numbers) or lagged (negative numbers) the aggregated country-level DHF/DSS waves. c) The association between timing of seasonal epidemics of a non-island regrouped ‘district’ and its distance to an urban center. An urban center is defined as the regrouped ‘district’ with the highest population density in a geographic region.

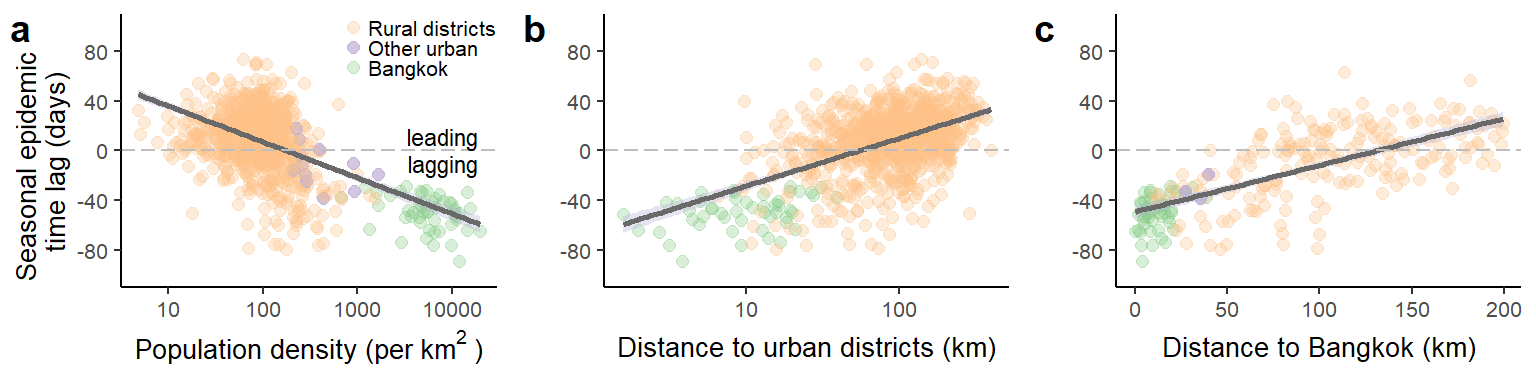

Figure S5: Recreation of Figure 2a-2c with all dengue cases, including cases of DF, DHF, and DSS. The figures show the associations of seasonal epidemic time lag with (a) the population density of a district, (b) the distance between a district and its regional urban center, defined as the district with the highest population density in a health region, and (c) the distance between central Bangkok to a district within a 200km radius from central Bangkok. The seasonal epidemic time lag is defined as the number of days the intra-annual dengue waves in a district preceded (positive numbers) or lagged (negative numbers) the aggregated country-level dengue waves. Each point represents a district, and colors represent the types of districts, including urban districts in Bangkok (green), other urban districts (defined as the districts on the mainland with the highest population density in each of the 12 health regions, not including Bangkok; purple), and all other districts (orange).

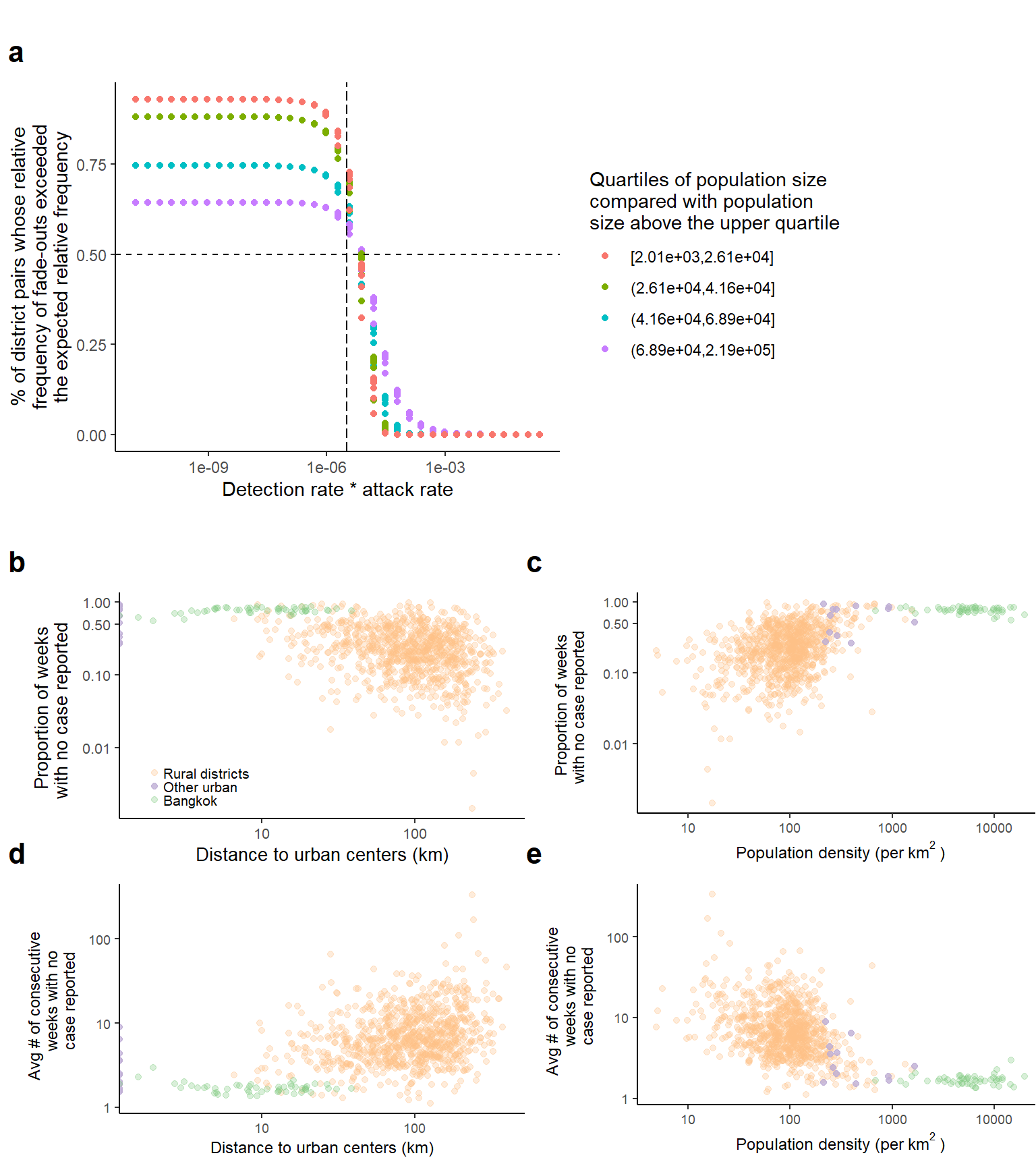

Figure S6: Comparing dengue fade-out between urban and rural districts. a) The proportion of district pairs whose relative frequency of dengue fade-outs exceeded the expected relative frequency based on sampling effects over a range of detection rate and attack rate. Here, we compared all districts with the districts whose population size fall above the upper quartile. Given the detection rate * attack rate estimated during the low dengue season (Jan-April) in Thailand (indicated by the dashed vertical line), the differences in fade-out frequency between districts were greater than would be expected based on sampling effects alone. b) The association between the proportion of weeks with no case reported over the study period and distance to urban centers. Each point represents a district, and colors represent 3 types of districts, districts in Bangkok (green), other urban districts (defined as the districts with the highest population density in each of the 12 health regions, not including Bangkok; purple), and all other districts (orange). c) The association between the proportion of weeks with no case reported over the study period and population density. d) The association between the average number of consecutive weeks with no case reported over the study period and distance to urban centers. e) The association between the average number of consecutive weeks with no case reported over the study period and population density.

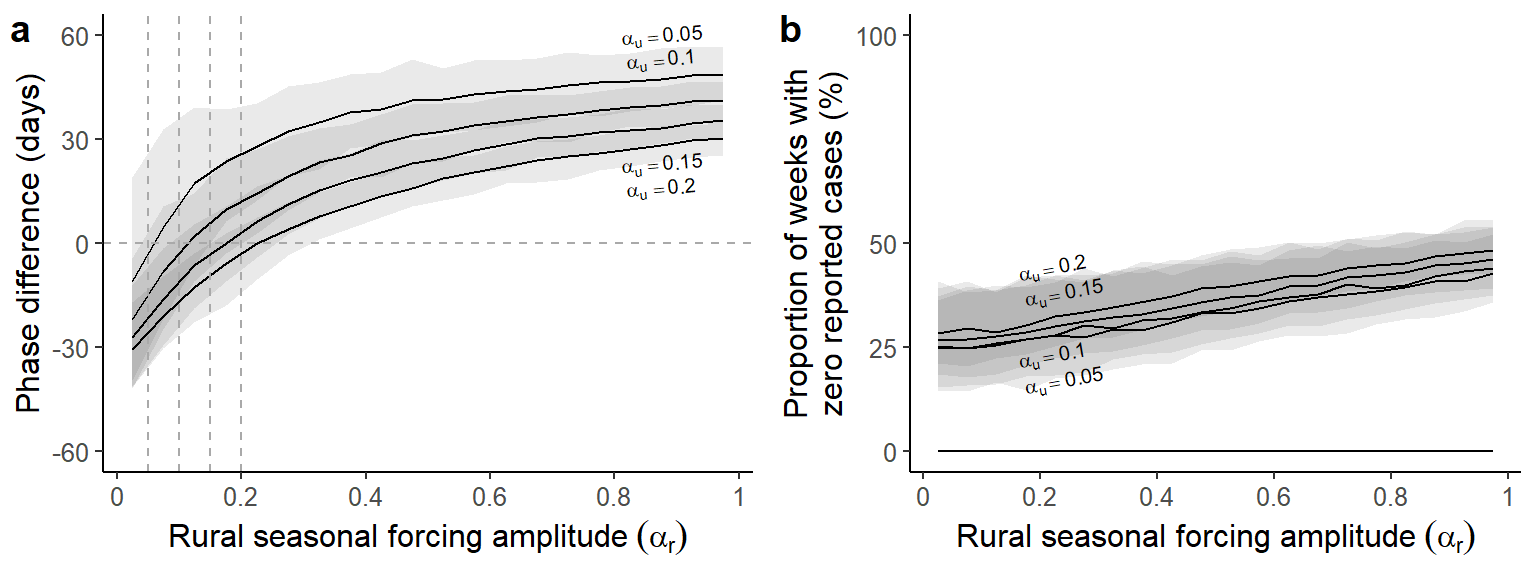

Figure S7: Relationship between timing of dengue seasonal epidemics, fadeout, and seasonal forcing in a simulated two-patch model. a) Phase differences of seasonal epidemics on the annual scale between the urban and rural patch for various parameter combinations. The parameters are: \(\alpha_{u}\)= 0.05, 0.10, 0.15, 0.2 (top to bottom), 0<\(\alpha_{r}\)<1, and \(m\) = 0.1. Positive phase difference indicates the number of days the intra-annual waves in the rural district preceded that in the urban patch, whereas negative phase difference indicates a lag. We ran 200 simulations for each combination of parameters for up to 40 years, and calculated average phase difference simulated seasonal epidemics (0.8 - 1.2 years) between the urban and rural patches. Solid black curves represent the smoothed median of the average phase differences. Shades represent the 95% confidence interval. b) the proportion of weeks without cases in simulated urban (dashed lines, all close to zero) and rural (solid lines) epidemics for the same set of parameters. \(\alpha_{u}\)= 0.05, 0.10, 0.15, 0.2 (bottom to top)

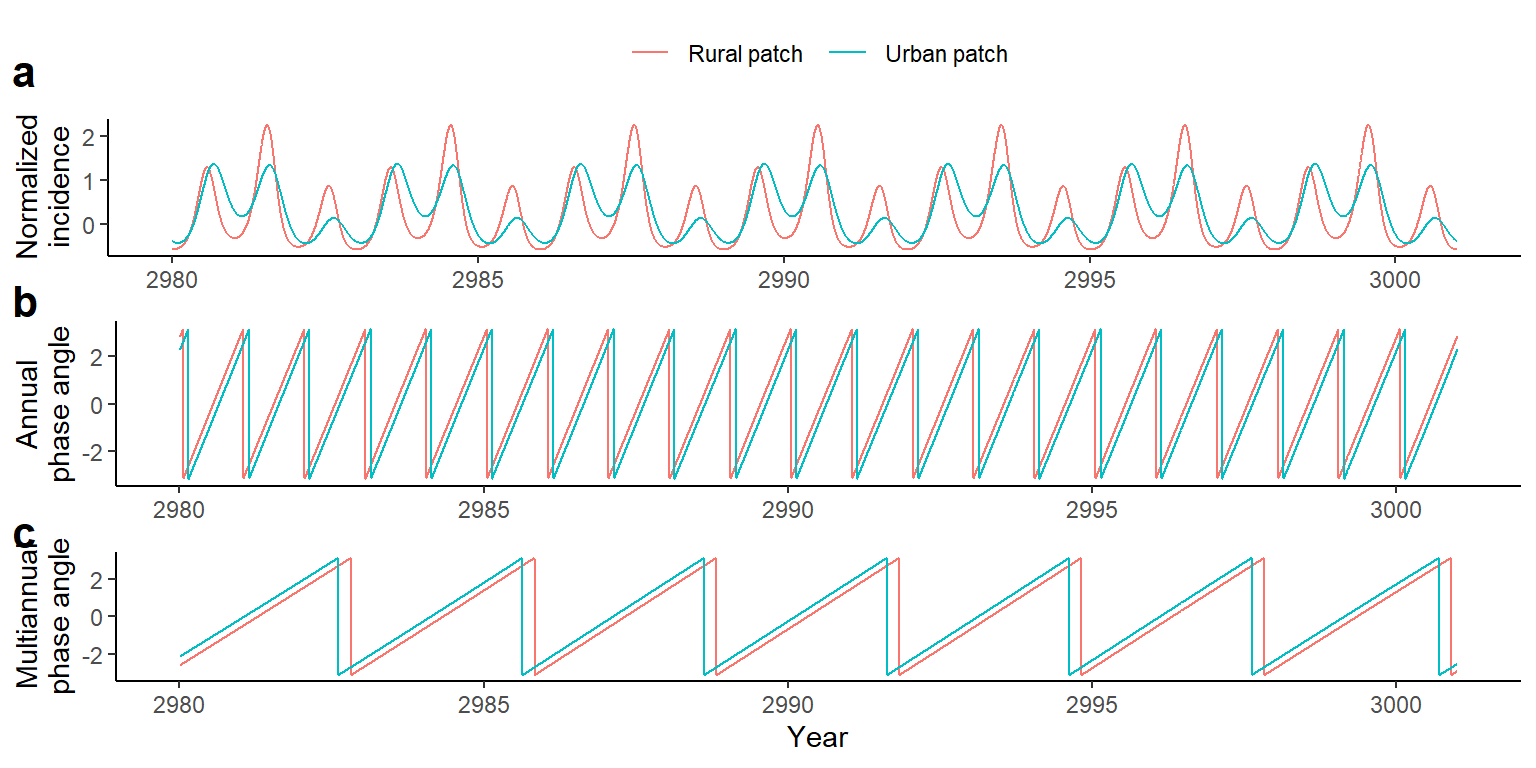

Figure S8: Theoretical consistency of urban patch leading multiannual waves while lagging seasonal waves. a) A 20-year time series of normalized incidence generated by the coupled SIR model with the updated rate of infection (See the section titled ‘Coupled Two-Patch SIR Models with Seasonal Forcing’ in the method for a detailed description of the method). Data for the urban patch were colored in blue, and rural patch were colored in red. b) Phase angle of the extracted annual (0.8-1.2 years) component of the normalized incidence. c) Phase angle of the extracted multiannual (2-5 years) component of the normalized incidence. On average, rural epidemics precede the urban epidemics by 31 days on the annual scale and lag the urban epidemics by 2.4 months on the multiannual scale. $\beta_{o}=0.15$, $\alpha_{u}=0.2$,$\alpha_{u}=0.8$, $m=0.1$.

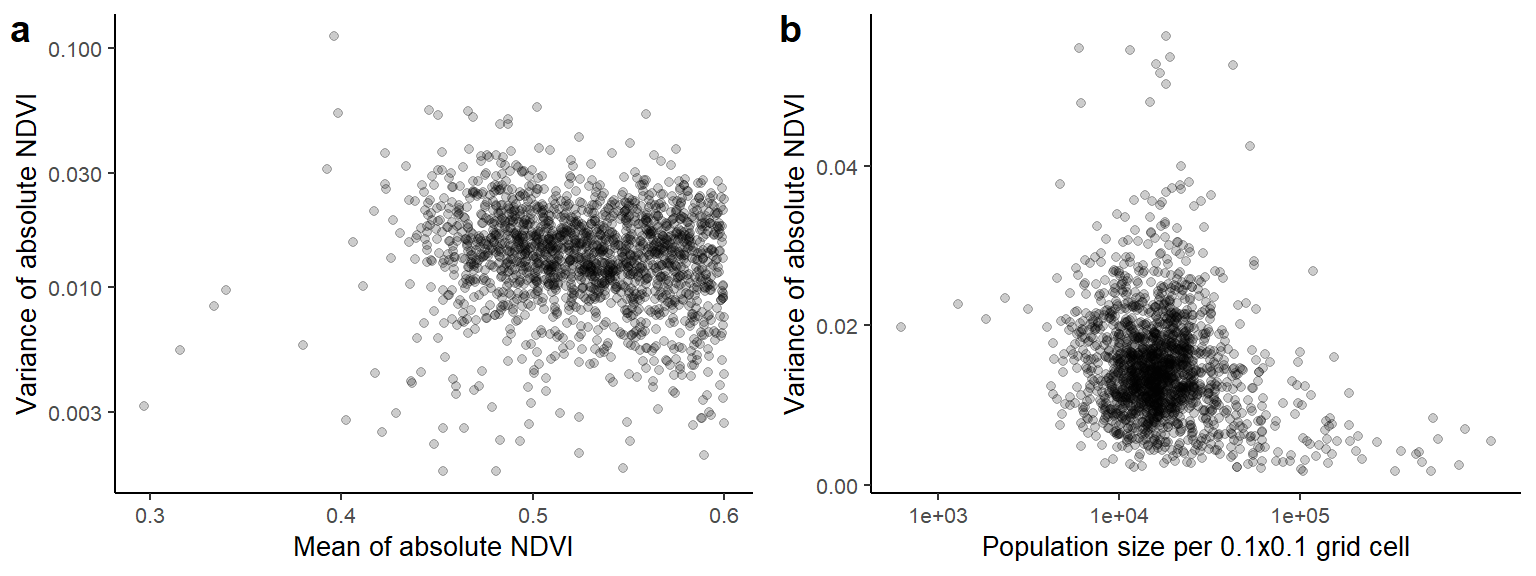

Figure S9: Seasonal variation of NDVI, mean NDVI, and population density. a) Association between variance of absolute NDVI and mean of absolute NDVI of each 0.1x0.1 grid cell in Thailand. b) Association between variance of absolute NDVI and population size of each 0.1x0.1 grid cell in Thailand. Data from tropical and temperate rainforests, herein defined as the grid cells with mean NDVI over 0.6, were excluded from both figures.

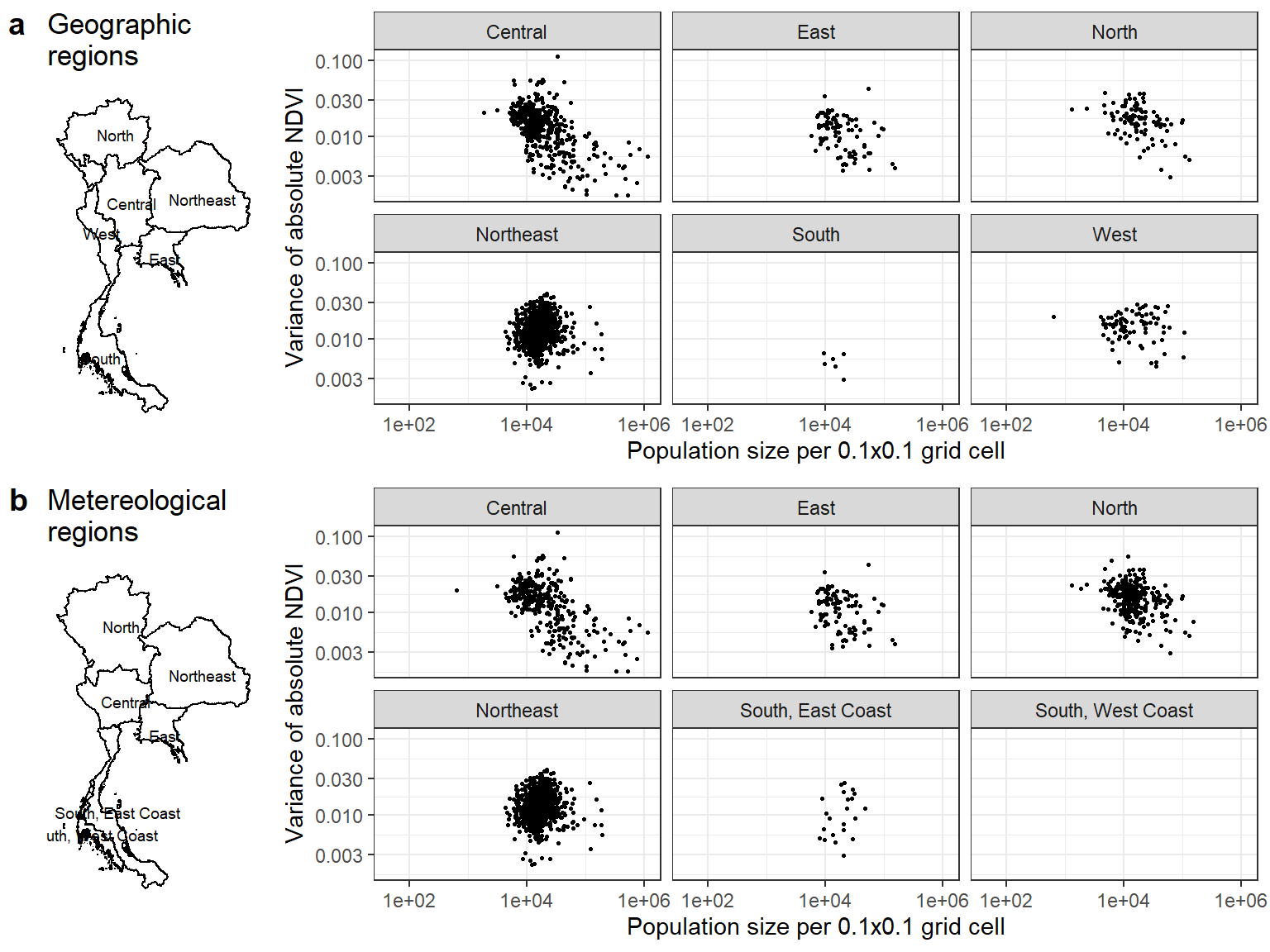

Figure S10: Association between variance of absolute NDVI and population size of each 0.1x0.1 grid cell by region, a) geographic region and b) meteorological region. Data from tropical and temperate rainforests, herein defined as the grid cells with mean NDVI over 0.6, were excluded.

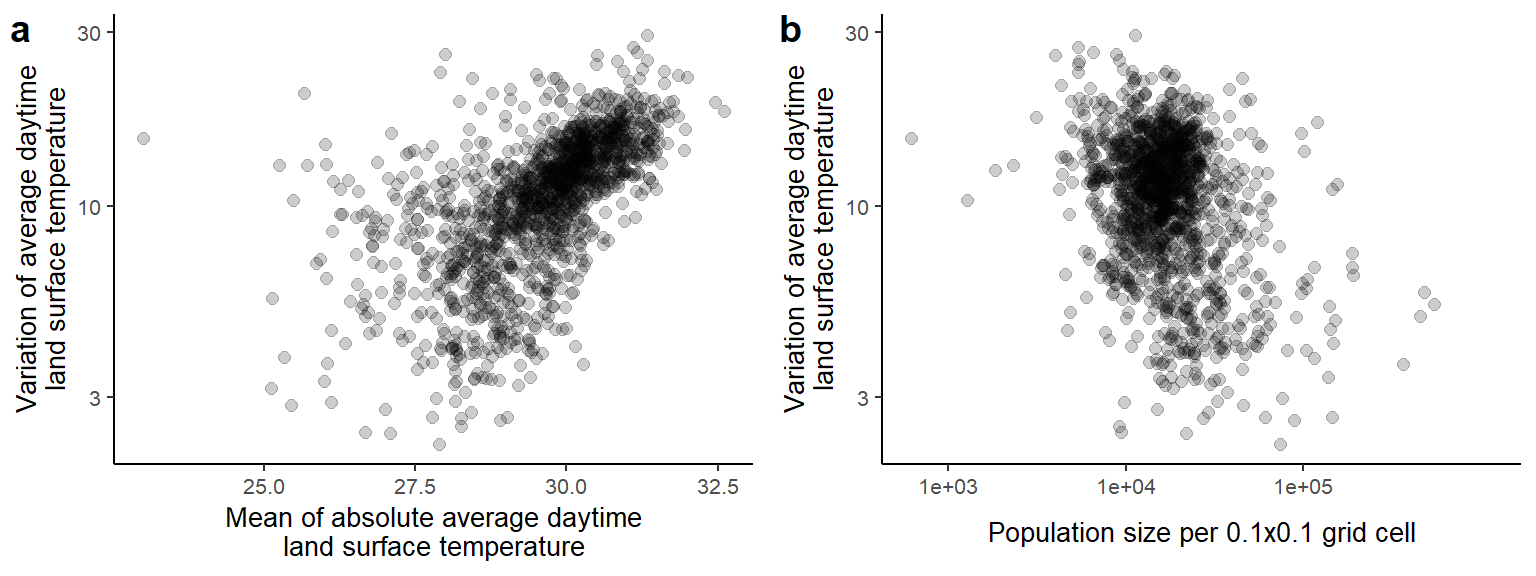

Figure S11: Seasonal variation and mean of daytime land surface temperature and population density. a) Association between variance and mean of absolute average daytime land surface temperature of each 0.1x0.1 grid cells in Thailand. b) Association between variance of absolute average daytime land surface temperature and population size of each 0.1x0.1 grid cells in Thailand. Data from tropical and temperate rainforests, herein defined as the grid cells with mean NDVI over 0.6, were excluded.

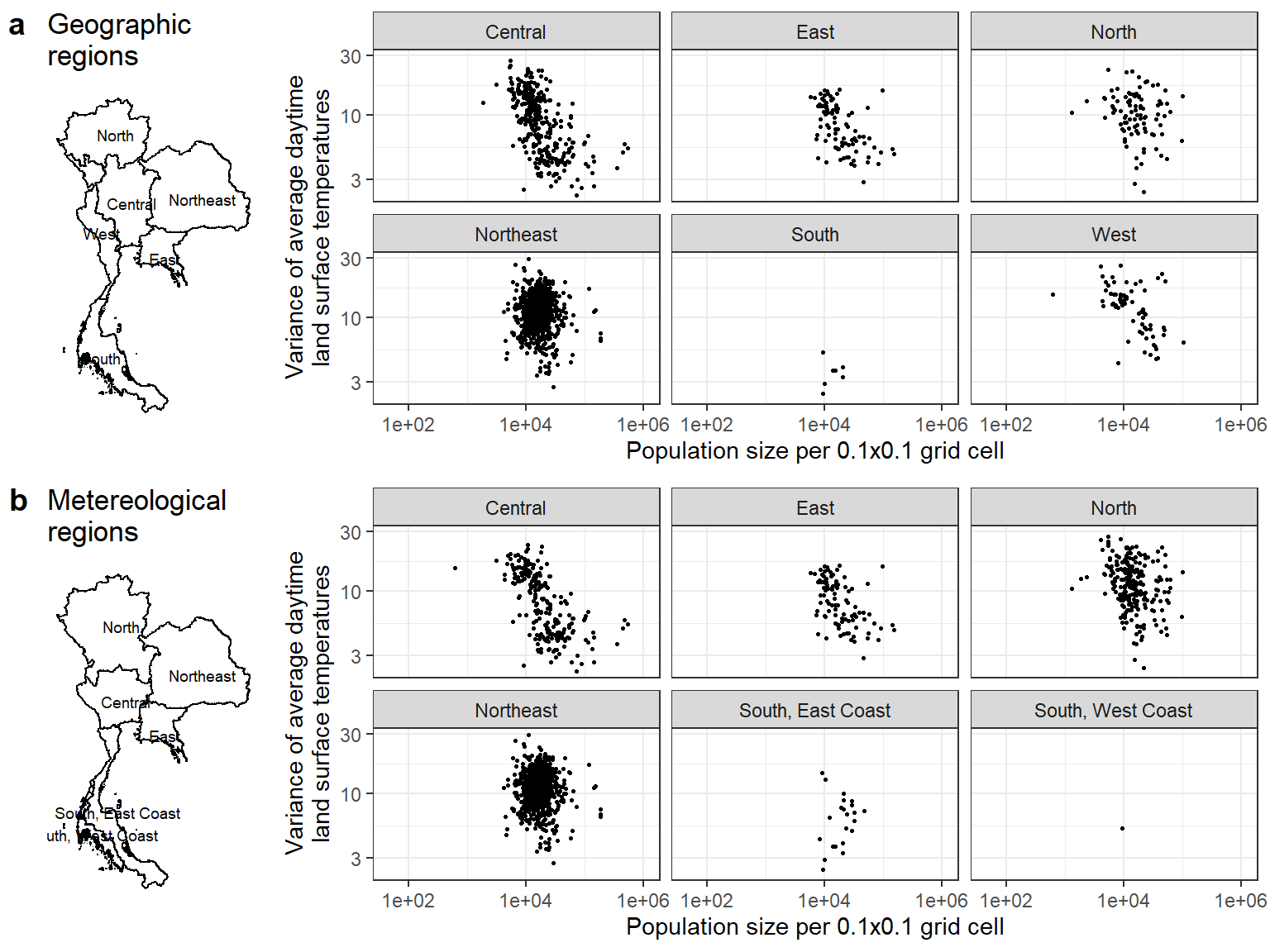

Figure S12: Association between variance of absolute average daytime land surface temperature and population size of each 0.1x0.1 grid cell by region, a) geographic region and b) meteorological region. Data from tropical and temperate rainforests, herein defined as the grid cells with mean NDVI over 0.6, were excluded.

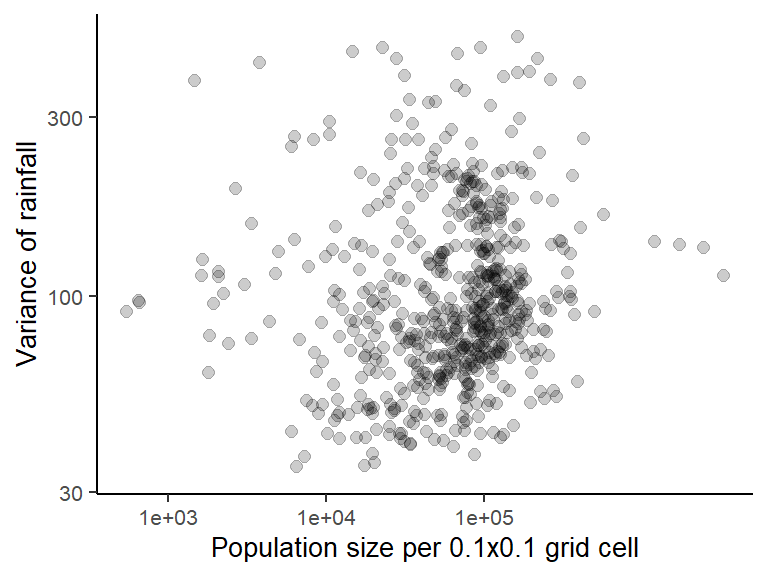

### Figure S13: Association between variance of rainfall and population size of each 0.25x0.25 grid cell in Thailand.

| Table S1.1. Relative contribution of population density and distance to urban centers for explaining the timing of seasonal dengue epidemics in each health region. Results indicate regression coefficients and 95% confidence interval of univariate and multivariate linear models. In univariate linear regression, predictors are either 1) population density on the logarithmic scale or 2) distance to urban centers. Multiple linear regression adjusted for both predictors. Observations from the regional urban centers are excluded from the model | | | | |
| --- | --- | --- | --- | --- |
|  | **Univariate linear regressions** | | **Bivariate linear regressions** | |
| **Health regions** | **Log population density** | **Distance to regional urban center * 10** | **Population density * log(2)** | **Distance to regional urban center * 10** |
| Region 1 | -0.66 [-4.1,2.78] | 0.63 [0.02,1.24] * | 0.14 [-3.31,3.59] | 0.63 [0.02,1.24] * |
| Region 2 | -1.27 [-7.78,5.25] | 0.67 [-0.59,1.93] | 0.58 [-6.73,7.89] | 0.72 [-0.69,2.14] |
| Region 3 | -13.12 [-21.86,-4.38] ** | 0.97 [-1.13,3.07] | -13.38 [-22.51,-4.25] ** | -0.18 [-2.37,2.02] |
| Region 4 | -8.08 [-12.17,-4] *** | 1.4 [0.05,2.75] * | -8 [-12.63,-3.37] *** | 0.05 [-1.48,1.59] |
| Region 5 | -14.42 [-17.9,-10.94] *** | 3.16 [2.39,3.93] *** | -8.94 [-13.38,-4.51] *** | 1.89 [0.91,2.87] *** |
| Region 6 | -8.5 [-12.48,-4.52] *** | 1.64 [0.98,2.31] *** | -4.03 [-8.92,0.87] | 1.24 [0.42,2.06] ** |
| Region 7 | 0.75 [-9.55,11.05] | 0.38 [-0.74,1.51] | 2.18 [-8.51,12.86] | 0.46 [-0.7,1.63] |
| Region 8 | 0.55 [-5.51,6.61] | -0.55 [-1.01,-0.08] * | -3.65 [-10.37,3.07] | -0.68 [-1.19,-0.17] ** |
| Region 9 | -2.41 [-9.22,4.4] | 0.99 [0.12,1.87] * | -4.48 [-11.32,2.37] | 1.13 [0.25,2.01] * |
| Region 10 | -3.8 [-12.5,4.9] | 0.39 [-0.65,1.43] | -2.98 [-14.82,8.86] | 0.14 [-1.27,1.55] |
| Region 11 | -8.11 [-13.01,-3.21] ** | 1.39 [0.74,2.03] *** | -4.35 [-9.66,0.95] | 1.14 [0.44,1.83] ** |
| Region 12 | -12.25 [-17.73,-6.77] *** | 1.24 [0.45,2.02] ** | -10.54 [-16.24,-4.84] *** | 0.71 [-0.11,1.53] |
| Bangkok | -4.06 [-9.8,1.68] | 6.17 [-0.8,13.14] | 1.7 [-9.95,13.36] | 7.98 [-6.17,22.13] |

| Table S1.2. Same as table 1.1 but the associations are assessed by geographic region | | | | | | | | |
| --- | --- | --- | --- | --- | --- | --- | --- | --- |
|  | | **Univariate linear regressions** | | | | **Bivariate linear regressions** | | |
| **Geographic regions** | | **Log population density** | | **Distance to regional urban center *10** | | **Population density * log(2)** | | **Distance to regional urban center * 10** |
| Central | | -8.85 [-9.94,-7.75] *** | | 1.9 [1.68,2.12] *** | | -4.85 [-6.34,-3.37] *** | | 1.15 [0.84,1.46] *** |
| East | | -3.92 [-9.65,1.8] | | 1.14 [0.3,1.99] ** | | -1.22 [-6.97,4.53] | | 1.07 [0.17,1.96] * |
| North | | -0.66 [-4.3,2.99] | | 0.63 [0.02,1.24] * | | 0.14 [-3.31,3.59] | | 0.63 [0.02,1.24] * |
| Northeast | | -3.91 [-7.6,-0.21] * | | 0.82 [0.59,1.05] *** | | -2.19 [-5.64,1.25] | | 0.8 [0.58,1.02] *** |
| South | | -14.73 [-18.28,-11.19] *** | | 1.37 [1.12,1.62] *** | | -7.08 [-10.95,-3.22] *** | | 1.08 [0.79,1.38] *** |
| West | | -5.18 [-9.42,-0.94] * | | 0.39 [-0.05,0.84] | | -4.5 [-8.95,-0.04] * | | 0.16 [-0.33,0.65] |

| Table S1.3. Same as table 1 but the associations are assessed by meteorological region | | | | |
| --- | --- | --- | --- | --- |
|  | **Univariate linear regressions** | | **Bivariate linear regressions** | |
| **Meteorological regions** | **Log population density** | **Distance to regional urban center * 10** | **Population density * log(2)** | **Distance to regional urban center * 10** |
| Central | -7.36 [-8.45,-6.28] *** | 3.06 [2.68,3.44] *** | -1.92 [-3.65,-0.2] * | 2.48 [1.84,3.12] *** |
| East | -2.53 [-7.93,2.87] | 1.14 [0.34,1.95] ** | 0.29 [-5.16,5.74] | 1.16 [0.3,2.03] ** |
| North | -1.3 [-4.21,1.6] | 0.39 [0.08,0.71] * | -1.56 [-4.28,1.15] | 0.41 [0.1,0.72] * |
| Northeast | -3.91 [-7.46,-0.35] * | 0.82 [0.6,1.04] *** | -2.19 [-5.54,1.16] | 0.8 [0.58,1.02] *** |
| South, East Coast | -12.12 [-15.51,-8.73] *** | 0.65 [0.48,0.82] *** | -8.3 [-11.9,-4.69] *** | 0.43 [0.24,0.63] *** |
| South, West Coast | -9.43 [-17.84,-1.01] * | -0.04 [-0.91,0.83] | -9.51 [-17.38,-1.64] * | -0.11 [-0.97,0.75] |

| Table S2.1. Relative contribution of distance to urban centers and distance to Bangkok for explaining the timing of dengue seasonal epidemics in each health region. Results indicate regression coefficients and 95% confidence interval of bivariate and multivariate linear models where phase difference (in units of days) is the outcome. Bivariate linear regression adjusted for both distance to urban centers and distance to Bangkok. Multiple linear regression adjusted for both distance to urban centers and distance to Bangkok in addition to population density. Observations from the regional urban centers are excluded from the model | | | |
| --- | --- | --- | --- |
|  | **Bivariate linear regression** | **Multivariate linear regressions** | |
| **Health regions** | **Distance to regional urban center * 10** | **Population density * log(2)** | **Distance to regional urban center * 10** |
| Region 1 | 0.77 [0.18,1.37] * | 1.23 [-2.17,4.62] | 0.83 [0.23,1.43] ** |
| Region 2 | 0.68 [-0.54,1.9] | 0.15 [-7.19,7.49] | 0.69 [-0.69,2.07] |
| Region 3 | 0.29 [-2.13,2.72] | -14.18 [-23.12,-5.24] ** | -1.18 [-3.75,1.38] |
| Region 4 | -5.07 [-7.76,-2.38] *** | 0.54 [-5.45,6.53] | -5.16 [-8,-2.32] *** |
| Region 5 | -8.72 [-14.12,-3.31] ** | -5.94 [-10.67,-1.22] * | -6.57 [-12.16,-0.99] * |
| Region 6 | -8.87 [-17.41,-0.33] * | -2.58 [-7.53,2.36] | -7.88 [-16.5,0.73] |
| Region 7 | 0.73 [-0.42,1.89] | 7.04 [-4.3,18.37] | 1.1 [-0.18,2.38] |
| Region 8 | -0.51 [-1.17,0.16] | -3.66 [-10.19,2.87] | -0.63 [-1.33,0.06] |
| Region 9 | 1.72 [0.04,3.4] * | -4.12 [-13.42,5.18] | 1.23 [-0.76,3.22] |
| Region 10 | 1.07 [-1.22,3.37] | -2.66 [-14.21,8.89] | 0.81 [-1.72,3.34] |
| Region 11 | 1.9 [1.12,2.67] *** | -4.83 [-10,0.34] | 1.66 [0.85,2.46] *** |
| Region 12 | 0.98 [0.03,1.93] * | -12.45 [-18.26,-6.64] *** | -0.02 [-1.07,1.02] |
| Bangkok | 6.17 [-0.55,12.89] | 1.7 [-9.62,13.03] | 7.98 [-5.77,21.73] |

| Table S2.2. Same as table 2.1 but the associations are assessed by geographic region | | | |
| --- | --- | --- | --- |
|  | **Bivariate linear regression** | **Multivariate linear regressions** | |
| **Geographic regions** | **Distance to regional urban center * 10** | **Population density * log(2)** | **Distance to regional urban center * 10** |
| Central | 1.9 [1.68,2.12] *** | -4.85 [-6.33,-3.38] *** | 1.15 [0.84,1.46] *** |
| East | 0.11 [-1.8,2.01] | -0.13 [-6.13,5.87] | 0.11 [-1.75,1.97] |
| North | 0.77 [0.16,1.38] * | 1.23 [-2.24,4.69] | 0.83 [0.21,1.44] ** |
| Northeast | 1.55 [0.51,2.59] ** | -1.17 [-5.23,2.89] | 1.36 [0.14,2.57] * |
| South | 1.76 [1.2,2.33] *** | -6.73 [-10.65,-2.81] *** | 1.31 [0.7,1.92] *** |
| West | -4.09 [-8.56,0.38] | -3.98 [-8.43,0.47] | -3.81 [-8.17,0.55] |

| Table S2.3. Same as table 2.1 but the associations are assessed by meteorological region | | | |
| --- | --- | --- | --- |
|  | **Bivariate linear regression** | **Multivariate linear regressions** | |
| **Meteorological regions** | **Distance to regional urban center * 10** | **Population density * log(2)** | **Distance to regional urban center * 10** |
| Central | 3.06 [2.68,3.44] *** | -1.92 [-3.65,-0.2] * | 2.48 [1.84,3.12] *** |
| East | 1.11 [-0.53,2.75] | 0.38 [-5.39,6.15] | 1.1 [-0.52,2.72] |
| North | 0.17 [-0.26,0.61] | -1.88 [-4.62,0.85] | 0.16 [-0.26,0.59] |
| Northeast | 1.55 [0.54,2.56] ** | -1.17 [-5.15,2.81] | 1.36 [0.17,2.55] * |
| South, East Coast | 0.67 [-0.13,1.47] | -10.3 [-14.32,-6.28] *** | -0.58 [-1.5,0.35] |
| South, West Coast | -0.01 [-0.89,0.86] | -10.49 [-20.48,-0.51] * | -0.13 [-0.99,0.73] |

| Table S3.1. Same analyses as those in Table S1.1 using a subset of data from districts where observed incidence pattern is consistent with results from wavelet analyses | | | | |
| --- | --- | --- | --- | --- |
|  | **Univariate linear regressions** | | **Bivariate linear regressions** | |
| **Health regions** | **Population density * log(2)** | **Distance to regional urban center * 10** | **Population density * log(2)** | **Distance to regional urban center * 10** |
| Region 1 | 1.64 [-5.49,8.77] | 0.35 [-1.04,1.73] | 2.13 [-4.96,9.22] | 0.44 [-0.94,1.81] |
| Region 2 | 0.83 [-14.1,15.75] | 0.95 [-1.41,3.3] | 2.28 [-12.57,17.14] | 1.03 [-1.31,3.36] |
| Region 3 | -15.87 [-30.32,-1.42] * | 2.44 [-0.97,5.86] | -14.04 [-29.7,1.62] | 0.97 [-2.73,4.66] |
| Region 4 | -8.93 [-14.46,-3.4] ** | 1.32 [-0.52,3.15] | -9.44 [-15.74,-3.13] ** | -0.32 [-2.4,1.77] |
| Region 5 | -14.92 [-19.28,-10.56] *** | 3.15 [2.18,4.12] *** | -9.82 [-15.47,-4.16] *** | 1.71 [0.45,2.96] ** |
| Region 6 | -9.14 [-14.92,-3.35] ** | 2.11 [1.18,3.04] *** | -3.42 [-9.9,3.06] | 1.84 [0.8,2.88] *** |
| Region 7 | -2.41 [-24.4,19.58] | -1.12 [-3.42,1.18] | -8.42 [-31.95,15.11] | -1.49 [-3.95,0.97] |
| Region 8 | -1.72 [-20.34,16.9] | -0.04 [-1.22,1.14] | -2.15 [-21.23,16.93] | -0.08 [-1.29,1.12] |
| Region 9 | -11.47 [-26.58,3.64] | 1.72 [0.14,3.3] * | -16.26 [-31.35,-1.18] * | 2.12 [0.54,3.7] ** |
| Region 10 | 3.59 [-12.43,19.61] | -0.01 [-2.11,2.1] | 10.91 [-16.35,38.17] | 1.17 [-2.4,4.73] |
| Region 11 | -11.21 [-19.81,-2.6] * | 1.02 [0.17,1.87] * | -8.42 [-17.55,0.71] | 0.68 [-0.22,1.59] |
| Region 12 | -14.3 [-22.17,-6.43] *** | 1.84 [0.79,2.9] *** | -10.64 [-18.86,-2.42] * | 1.32 [0.23,2.42] * |
| Bangkok | -4.76 [-11.09,1.57] | 6.85 [-0.78,14.47] | 1.24 [-11.86,14.34] | 8.16 [-7.59,23.92] |

| Table S3.2. Same analyses as those in Table S1.2 using a subset of data from districts where observed incidence pattern is consistent with results from wavelet analyses | | | | |
| --- | --- | --- | --- | --- |
|  | **Univariate linear regressions** | | **Bivariate linear regressions** | |
| **Geographic regions** | **Population density * log(2)** | **Distance to regional urban center * 10** | **Population density * log(2)** | **Distance to regional urban center * 10** |
| Central | -8.56 [-9.98,-7.14] *** | 2.1 [1.81,2.4] *** | -3.47 [-5.46,-1.48] *** | 1.5 [1.04,1.95] *** |
| East | -4.61 [-12.55,3.33] | 1.76 [0.65,2.86] ** | -1.34 [-8.74,6.05] | 1.69 [0.56,2.83] ** |
| North | 1.64 [-6.22,9.49] | 0.35 [-1.04,1.74] | 2.13 [-5.03,9.29] | 0.44 [-0.95,1.83] |
| Northeast | -8.46 [-17.15,0.23] | 1.11 [0.65,1.57] *** | -5.45 [-13.29,2.39] | 1.06 [0.6,1.51] *** |
| South | -21 [-26.73,-15.27] *** | 1.69 [1.35,2.02] *** | -9.56 [-15.63,-3.49] ** | 1.35 [0.96,1.74] *** |
| West | -5.88 [-12.02,0.26] | 0.3 [-0.45,1.05] | -6.03 [-12.07,0.02] | -0.05 [-0.86,0.77] |

| Table S3.3. Same analyses as those in Table S1.3 using a subset of data from districts where observed incidence pattern is consistent with results from wavelet analyses | | | | |
| --- | --- | --- | --- | --- |
|  | **Univariate linear regressions** | | **Bivariate linear regressions** | |
| **Meteorological regions** | **Population density * log(2)** | **Distance to regional urban center * 10** | **Population density * log(2)** | **Distance to regional urban center * 10** |
| Central | -6.4 [-7.86,-4.94] *** | 2.83 [2.31,3.36] *** | -1.06 [-3.4,1.29] | 2.5 [1.6,3.4] *** |
| East | -4.35 [-11.94,3.25] | 1.73 [0.64,2.83] ** | -1.12 [-8.43,6.18] | 1.68 [0.55,2.81] ** |
| North | 0.52 [-5.72,6.75] | 0.82 [0.32,1.32] ** | -0.54 [-6.3,5.23] | 0.83 [0.33,1.32] ** |
| Northeast | -8.46 [-16.84,-0.09] * | 1.11 [0.66,1.56] *** | -5.45 [-13.26,2.35] | 1.06 [0.61,1.51] *** |
| South, East Coast | -15.69 [-20.86,-10.52] *** | 0.76 [0.52,0.99] *** | -10.42 [-15.91,-4.92] *** | 0.5 [0.24,0.77] *** |
| South, West Coast | -10.82 [-22.43,0.78] | 0.29 [-0.81,1.38] | -10.65 [-21.36,0.06] | 0.19 [-0.89,1.27] |

| Table S4.1. Same analyses as those in Table S2.1 using a subset of data from districts where observed incidence pattern is consistent with results from wavelet analyses | | | |
| --- | --- | --- | --- |
|  | **Bivariate linear regression** | **Multivariate linear regressions** | |
| **Health regions** | **Distance to regional urban center * 10** | **Population density * log(2)** | **Distance to regional urban center * 10** |
| Region 1 | 0.73 [-0.74,2.21] | 1.35 [-5.77,8.47] | 0.77 [-0.7,2.24] |
| Region 2 | 1.03 [-1.5,3.55] | 4.39 [-13.59,22.38] | 1.37 [-1.49,4.22] |
| Region 3 | 1.88 [-1.76,5.52] | -15.77 [-31.52,-0.02] | -0.08 [-4.17,4.01] |
| Region 4 | -4.04 [-7.31,-0.77] * | -0.11 [-9.76,9.53] | -4.02 [-7.6,-0.44] * |
| Region 5 | -10.48 [-18.08,-2.88] ** | -7.39 [-13.25,-1.54] * | -8.55 [-16.21,-0.9] * |
| Region 6 | -2.45 [-14.13,9.24] | -2.95 [-9.59,3.7] | -1.23 [-13.09,10.63] |
| Region 7 | -0.06 [-2.51,2.38] | -1.93 [-26.02,22.17] | -0.17 [-2.93,2.59] |
| Region 8 | -0.03 [-1.8,1.75] | -2.24 [-21.69,17.21] | -0.11 [-2.02,1.79] |
| Region 9 | 4.5 [1.13,7.87] ** | -12.84 [-34.08,8.39] | 2.99 [-1.17,7.15] |
| Region 10 | 3.23 [-1.68,8.13] | 8.78 [-18.28,35.85] | 4.01 [-1.4,9.42] |
| Region 11 | 1.18 [0.09,2.28] * | -9.52 [-18.81,-0.23] * | 0.99 [-0.11,2.09] |
| Region 12 | 2.16 [0.85,3.46] ** | -10.94 [-19.63,-2.25] * | 1.22 [-0.27,2.71] |
| Bangkok | 6.85 [-0.53,14.22] | 1.24 [-11.68,14.17] | 8.16 [-7.38,23.7] |

| Table S4.2. Same analyses as those in Table S2.2 using a subset of data from districts where observed incidence pattern is consistent with results from wavelet analyses | | | |
| --- | --- | --- | --- |
|  | **Bivariate linear regression** | **Multivariate linear regressions** | |
| **Geographic regions** | **Distance to regional urban center * 10** | **Population density * log(2)** | **Distance to regional urban center * 10** |
| Central | 2.1 [1.81,2.39] *** | -3.47 [-5.44,-1.5] *** | 1.5 [1.05,1.94] *** |
| East | 0.27 [-2.22,2.76] | -0.09 [-7.67,7.49] | 0.27 [-2.18,2.72] |
| North | 0.73 [-0.78,2.25] | 1.35 [-5.89,8.58] | 0.77 [-0.73,2.26] |
| Northeast | 4.15 [1.89,6.4] *** | -0.84 [-9.49,7.81] | 4.04 [1.55,6.52] ** |
| South | 2.22 [1.42,3.02] *** | -9.08 [-15.18,-2.98] ** | 1.74 [0.89,2.59] *** |
| West | -5.31 [-11.83,1.2] | -5.26 [-11.35,0.84] | -4.64 [-11.07,1.79] |

| Table S4.3. Same analyses as those in Table S2.3 using a subset of data from districts where observed incidence pattern are consistent with results from wavelet analyses | | | |
| --- | --- | --- | --- |
|  | **Bivariate linear regression** | **Multivariate linear regressions** | |
| **Meteorological regions** | **Distance to regional urban center * 10** | **Population density * log(2)** | **Distance to regional urban center * 10** |
| Central | 2.83 [2.31,3.35] *** | -1.06 [-3.39,1.27] | 2.5 [1.61,3.39] *** |
| East | 0.77 [-1.48,3.02] | -0.14 [-7.7,7.43] | 0.77 [-1.46,3] |
| North | 0.35 [-0.48,1.19] | -1.84 [-7.82,4.15] | 0.31 [-0.53,1.15] |
| Northeast | 4.15 [1.91,6.38] *** | -0.84 [-9.47,7.79] | 4.04 [1.56,6.51] ** |
| South, East Coast | 1.46 [0.22,2.71] * | -11.16 [-17.25,-5.08] *** | 0.11 [-1.32,1.54] |
| South, West Coast | 0.29 [-0.8,1.38] | -12.58 [-26.28,1.12] | 0.17 [-0.91,1.25] |

| Table S5. Impact of variation in plausible climatic drivers on timing of seasonal dengue epidemics. The results indicate the coefficients and 95% CI of univariate and bivariate linear regression models where phase difference (in units of days) is the outcome and population density on logarithmic scale and variance of climatic factors are the explanatory variables. | | |
| --- | --- | --- |
| **Variables** | **Univariate linear regression: coefficient (95% CI)** | **Bivariate linear regression: coefficient (95% CI)** |
| **1) Impact of variation in NDVI** | | |
| Variance of NDVI | 370.28 [187.59,552.96] *** | 230.64 [-45.92,507.2] |
| Population density * log(2) |  | -9.49 [-10.52,-8.45] *** |
| **2) Impact of variation in land surface temperature** | | |
| Variance of daytime land surface temperature | 0.78 [0.51,1.05] *** | 1.33 [0.88,1.79] *** |
| Population density * log(2) |  | -7.2 [-8.69,-5.72] *** |
| **3) Impact of variation in rainfall** | | |
| Variance of rainfall | -0.01 [-0.03,0] | 0 [-0.02,0.02] |
| Population density * log(2) |  | -9.08 [-9.96,-8.19] *** |
